# A Comprehensive Statistical Analysis of COVID-19 Trends: Global and U.S. Insights through ARIMA, Regression, and Spatial Models

**DOI:** 10.1101/2024.10.22.24315932

**Authors:** Zhihao Lei

## Abstract

The COVID-19 pandemic has driven the need for accurate data analysis and forecasting to guide public health decisions. In this study, we utilized ARIMA and ARIMAX models to predict short-term trends in confirmed COVID-19 cases across different regions, including the United States, Asia, Europe, Africa, and the Americas. Comparisons were made between ARIMA and auto.arima models, and anomaly detection was performed to investigate discrepancies between predictions and actual data. The study also explored the relationship between vaccination rates and new case numbers, and examined how socioeconomic factors such as GDP per capita, HDI, and healthcare resources influenced COVID-19 incidence rates across countries. Our findings provide insights into the effectiveness of predictive models and the significant impact of socioeconomic factors on the spread of the virus, contributing valuable information for future epidemic prevention and control strategies.

## 1 INTRODUCTION

Since the onset of the COVID-19 pandemic in late 2019, the virus has had profound and widespread effects on public health, economies, and daily life across the globe. As of 2024, the pandemic continues to challenge healthcare systems, and accurate forecasting of COVID-19 case trends remains critical for effective policy-making and intervention strategies. Statistical modeling, particularly time series analysis, has proven to be a valuable tool in predicting the trajectory of the pandemic and assisting in the formulation of public health responses (Lai, Shih, Ko, Tang, & Hsueh, 2020).

Among the various statistical models, the AutoRegressive Integrated Moving Average (ARIMA) model has been widely employed in epidemiological studies for short-term forecasting due to its simplicity and effectiveness in modeling temporal data (Adhikari & Agrawal, 2013). ARIMA models have been used to predict COVID-19 case trends in various countries, demonstrating that these models can provide reasonably accurate forecasts over short time horizons (Benvenuto, Giovanetti, Vassallo, Angeletti, & Ciccozzi, 2020). However, the accuracy of ARIMA-based forecasts can vary significantly across regions and time periods due to factors such as virus mutations, government interventions, and changes in population behavior (Petropoulos & Makridakis, 2020). One notable limitation of ARIMA models is their reliance on historical data alone, without considering external factors that might influence future trends, such as vaccination rates, policy shifts, or behavioral changes. This limitation can result in higher uncertainty when making long-term predictions.

To address these limitations, the AutoRegressive Integrated Moving Average with Exogenous Variables (ARIMAX) model introduces external variables, such as vaccination rates, to enhance the predictive power of the model. Incorporating vaccination data allows researchers to assess the potential impact of vaccination campaigns on future case numbers, offering a more comprehensive view of epidemic dynamics (Bontempi, Vergalli, & Squazzoni, 2021). While previous studies have shown that vaccination plays a crucial role in mitigating the spread of COVID-19, leading to significant reductions in new case numbers following large-scale immunization efforts (Paltiel, Zheng, & Schwartz, 2021), most existing research has focused on specific regions or time periods, lacking a holistic analysis of the complex interactions between vaccination, virus mutations, and policy interventions.

In addition to time series forecasting, understanding the relationship between COVID-19 incidence and socioeconomic factors is crucial. Previous research has highlighted the role of GDP per capita, healthcare infrastructure, and other socioeconomic indicators in shaping the pandemic’s impact across different regions (Islam, Khunti, Dambha-Miller, Kawachi, & Marmot, 2021). For example, countries with higher healthcare spending and better medical resources have been better equipped to manage the crisis, resulting in lower mortality rates and more effective containment strategies (Bambra, Riordan, Ford, & Matthews, 2020). However, many studies are limited to single-variable analyses and fail to fully account for the multifaceted interactions among these socioeconomic factors, which can contribute to significant disparities in COVID-19 outcomes across different countries.

This study seeks to advance the existing body of work by applying both ARIMA and ARIMAX models to predict short-term COVID-19 case trends in the United States and globally. By introducing vaccination rates as an exogenous variable in the ARIMAX model, we aim to improve the accuracy of predictions and provide deeper insights into the dynamics between vaccination efforts and new case trends. Moreover, by analyzing discrepancies between predicted and actual case numbers, we investigate potential causes for forecast anomalies, such as policy shifts and virus mutations. Additionally, we examine how socioeconomic factors—including GDP per capita, healthcare resources, and the Human Development Index (HDI)—influence COVID-19 incidence rates across countries, offering a more comprehensive understanding of the pandemic’s broader determinants. Through this multidimensional approach, our study not only compares the performance of ARIMA and ARIMAX models but also contributes valuable insights into the factors driving the spread of COVID-19, thereby informing future epidemic prevention and control strategies.

## 2 DATA COLLECTION

To conduct a comprehensive analysis of the COVID-19 pandemic and its associated factors, a diverse range of datasets was selected from reputable sources such as the World Health Organization (WHO), Centers for Disease Control and Prevention (CDC), World Bank, and other national and international agencies. These datasets were carefully chosen based on their relevance, comprehensiveness, and frequency of updates, ensuring that the analysis reflects the most accurate and up-to-date information available. As shown in Table 1, the datasets include daily and weekly reported COVID-19 cases, deaths, and vaccination trends, along with key socioeconomic indicators such as GDP per capita, Human Development Index (HDI), Gini index, healthcare expenditures, and healthcare infrastructure data. These variables were critical for modeling and understanding the progression of the pandemic and the impact of various factors on infection rates.

**TABLE 1.** Overview of Key Datasets.

| Data Source | Data Description | Link |
| --- | --- | --- |
| WHO | Daily COVID-19 cases and deaths reported to WHO. Updated weekly. Includes corrections to historical data based on additional information received. | https://data.who.int/dashboards/covid19/data?n=c |
| CDC(Centers for Disease Control and Prevention) | COVID-19 Vaccination Trends in the United States at national and jurisdictional levels. Updated regularly with trends over time. | https://data.cdc.gov/Vaccinations/COVID-19-Vaccination-Trends-in-the-United-States-N/rh2h-3yt2/about_data |
| World Bank | GDP per capita (current US\$), representing the monetary value of all finished goods and services per person. | https://data.worldbank.org/indicator/NY.GDP.PCAP.CD |
| UNDP(United Nations Development Programme) | Human Development Index (HDI), measuring key dimensions of human development: a long and healthy life, access to education, and a decent standard of living. | https://hdr.undp.org/data-center/human-development-index#/indicies/HDI |
| World Bank | Gini index, measuring the distribution of income across a population, representing inequality. | https://data.worldbank.org/indicator/SI.POV.GINI |
| World Bank | Current health expenditure per capita (current US\$), reflecting national spending on health. | https://data.worldbank.org/indicator/SH.XPD.CHEX.PC.CD |
| World Bank | Hospital beds per 1,000 people, indicating the availability of healthcare infrastructure. | https://data.worldbank.org/indicator/SH.MED.BEDS.ZS |
| World Bank | Population density (people per sq. km of land area), representing the concentration of people in a given area. | https://data.worldbank.org/indicator/EN.POP.DNST |
| CDC(Centers for Disease Control and Prevention) | Weekly United States COVID-19 Cases and Deaths by State - ARCHIVED data. Includes state-level trends and historical data. | https://data.cdc.gov/Case-Surveillance/Weekly-United-States-COVID-19-Cases-and-Deaths-by-/pwn4-m3yp/about_data |
| CDC(Centers for Disease Control and Prevention) | COVID-19 Vaccinations in the United States at the county level, with detailed trends and demographic breakdowns. | https://data.cdc.gov/Vaccinations/COVID-19-Vaccinations-in-the-United-States-County/8xxk-amqh/about_data |
| BEA(Bureau of Economic Analysis) | Gross Domestic Product by State and Personal Income by State for the 1st Quarter of 2024, offering insights into regional economic performance. | https://www.bea.gov/data/gdp/gdp-state |
| U.S. Census Bureau | Historical Population Density Data (1910-2020), providing population density trends over the past century. | https://www.census.gov/data/tables/time-series/dec/density-data-text.html |
| U.S. Census Bureau | State Population Totals and Components of Change from 2020-2023, showing population changes and migration trends. | https://www.census.gov/data/tables/time-series/demo/popest/2020s-state-total.html |

TABLE 1 Overview of Key Datasets: continued
| Data Source | Data Description | Link |
| --- | --- | --- |
| Kaiser Family Foundation (KFF) | Health Care Expenditures per Capita by State of Residence, detailing state-level spending on healthcare. | https://www.kff.org/other/state-indicator/health-spending-per-capita/?currentTimeframe=0&sortModel=%7B%22colId%22:%22Location%22,%22sort%22:%22asc%22%7D |
| Kaiser Family Foundation (KFF) | Hospital Beds per 1,000 Population by Ownership Type, offering insights into the distribution of healthcare resources. | https://www.kff.org/other/state-indicator/beds-by-ownership/?currentTimeframe=0&sortModel=%7B%22colId%22:%22Location%22,%22sort%22:%22asc%22%7D |

In this paper, all analyses were performed using R.

## 3 METHODOLOGY

### 3.1 Theoretical Basis of the ARIMA model

The ARIMA model is a widely utilized statistical method for analyzing and forecasting time series data. Its general form for an ARIMA model of order (*p, d, q*) is given by the following equation:

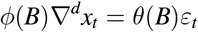

Where:

- ∇^*d*^ = (1–*B*)^*d*^ is the differencing operator, with *B* representing the backshift operator (Box, Jenkins, Reinsel, & Ljung, 2015).
- *ϕ*(*B*) = 1 – *ϕ*_1_*B* – …– *ϕ*_*p*_*B*^*p*^ is the autoregressive (AR) coefficient polynomial (Box et al., 2015).
- *θ*(*B*) = 1 – *θ*_1_*B* – …– *θ*_*q*_*B*^*q*^ is the moving average (MA) coefficient polynomial (Box et al., 2015).
- *ε*_*t*_ denotes the white noise error terms, which satisfy the following properties: *E* (*ε*_*t*_) = 0, 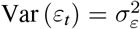, and *E* (*ε*_*t*_*ε*_*s*_) = 0 for *s* ≠ *t* (Box et al., 2015).
- Furthermore, *E* (*x*_*s*_*ε*_*t*_) = 0 for all *s* < *t*, ensuring that the noise terms are uncorrelated with past values of the series (Hamilton, 1994).

The ARIMA model is composed of three primary components:

1. Autoregressive (AR): The AR part represents the dependence between an observation and several lagged observations (Box et al., 2015).
2. Integrated (I): The integrated component represents the differencing needed to make the time series stationary (Box et al., 2015).
3. Moving Average (MA): The MA part models the dependency between an observation and residual errors from a moving average model applied to lagged observations (Box et al., 2015).

The ARIMA modeling process begins with a stationarity test, commonly performed using the Augmented Dickey-Fuller (ADF) test (Dickey & Fuller, 1979). If the time series is non-stationary, transformations such as differencing or logarithmic scaling are applied to achieve stationarity (Hamilton, 1994). Next, model identification involves determining the order of the model, specifically the values of *p* and *q*, which are the autoregressive and moving average terms, respectively. This is usually done by analyzing the autocorrelation function (ACF) and partial autocorrelation function (PACF) plots (Box et al., 2015). The differencing order *d* is selected based on the transformations applied during the stationarity phase. After the model is identified, the parameters *ϕ*_*i*_ and *θ*_*j*_ are estimated, typically using maximum likelihood estimation (MLE) (Hamilton, 1994). Model validation then follows, where statistical tests such as the Ljung-Box test are used to ensure the residuals behave like white noise, indicating that the model has adequately captured the time series’ structure (Ljung & Box, 1978). Model selection is based on criteria like the Akaike Information Criterion (AIC) or Bayesian Information Criterion (BIC), with the model having the lowest criterion value generally preferred (Akaike, 1974). Finally, once validated, the model is used for forecasting future values of the time series (Box et al., 2015).

The process is visually represented in Figure 1, which illustrates the steps of fitting an ARIMA model to a time series, starting from stationarity checks to forecasting.

**FIGURE 1.**
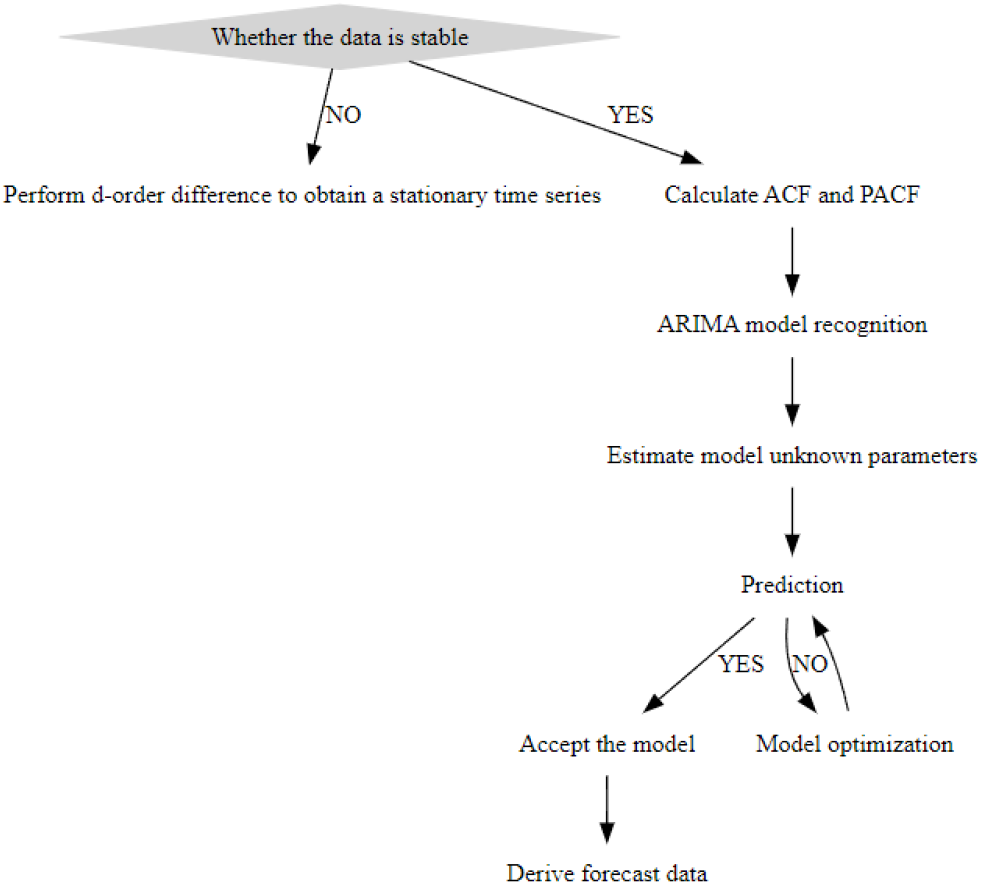
ARIMA Model Construction Flow Chart

### 3.2 Rolling Window Cross-Validation and Comparison with auto.arima

In this study, rolling window cross-validation was used to evaluate the performance of ARIMA models for time series forecasting. The primary goal was to identify the optimal ARIMA model parameters by minimizing the Root Mean Squared Error (RMSE) and to compare the results with those obtained from the automated model selection function, `auto.arima` (Hyndman & Athanasopoulos, 2018).

Rolling window cross-validation is a method specifically designed for time series data, which preserves the temporal order of the observations. In each iteration, a model is trained on a fixed-length window of historical data and then validated on the subsequent observation. This approach ensures that the evaluation reflects real-world forecasting scenarios, where future values are predicted based on past data (Bergmeir & Benítez, 2012). For each ARIMA model evaluated, the one-step-ahead forecast errors were calculated, and RMSE was used as the evaluation metric. RMSE is given by:

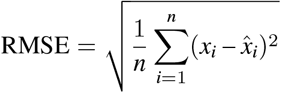

where *x*_*i*_ are the actual values and 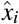 are the predicted values. Lower RMSE values indicate better model performance (Chai & Draxler, 2014). A grid search was performed over various combinations of autoregressive (*p*) and moving average (*q*) parameters, with the differencing order (*d*) fixed at 1. This process was parallelized to efficiently explore the parameter space (Hyndman & Athanasopoulos, 2018).

The reason for selecting RMSE as the evaluation metric lies in its ability to quantify the average prediction error while giving more weight to larger errors (Willmott & Matsuura, 2005). RMSE is particularly useful in contexts where significant deviations in the forecast could have a considerable impact, as it highlights larger discrepancies between predicted and actual values more than other metrics, such as Mean Absolute Error (MAE). Additionally, RMSE is measured in the same units as the original data, making the results easier to interpret in practical applications.

To compare the performance of manual model selection with automated methods, the `auto.arima` function was used. `auto.arima` automatically selects the best ARIMA model by optimizing information criteria such as the Akaike Information Criterion (AIC) or Bayesian Information Criterion (BIC) (Hyndman & Khandakar, 2008). While `auto.arima` quickly identifies a suitable model based on a global fit to the entire dataset, rolling window cross-validation provides a more dynamic evaluation by assessing the model’s performance across different periods in the data (Tashman, 2000). This allows for a comparison of how well the automated model selection aligns with the results of cross-validated manual tuning.

By visualizing the RMSE values across different parameter combinations, we were able to compare the best-performing model identified by rolling window cross-validation with the model chosen by `auto.arima`. This comparison provided insights into the trade-offs between automated and manual ARIMA model selection in time series forecasting.

## 3.3 Anomaly Detection

Anomaly detection in time series data is crucial for identifying irregular patterns, such as sudden spikes in COVID-19 case counts. In this study, we employed a statistical approach to detect anomalies directly within the time series data without explicitly fitting a complex model like ARIMA. This approach is commonly referred to as residual-based anomaly detection, where outliers are identified based on their deviation from expected patterns in the residuals (Hyndman & Athanasopoulos, 2018).

The anomaly detection approach used in this study is grounded in statistical rules that flag data points as anomalies when they significantly deviate from surrounding values. Specifically, the detection mechanism works by identifying outliers in the residuals after accounting for typical patterns in the time series (Chandola, Banerjee, & Kumar, 2009). These residuals are examined, and points are flagged as outliers if they exceed a certain threshold of deviation from the local mean.

The theoretical basis for this method relies on identifying points that deviate significantly from the local mean or expected value of the time series. In mathematical terms, a data point *x*_*t*_ is considered an outlier if it satisfies:

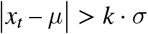

where *µ* represents the local mean, *σ* is the standard deviation of the surrounding data points, and *k* is a threshold factor that determines the sensitivity of the detection (Aggarwal, 2017). Commonly, *k* is set to values such as 2 or 3, which correspond to confidence intervals typically used in outlier detection (Chandola et al., 2009).

This method is particularly effective for detecting additive outliers, which manifest as sudden spikes or drops in the series, such as those that might occur due to external shocks like the emergence of a new COVID-19 variant (Hyndman & Athanasopoulos, 2018). By identifying and analyzing these outliers, we can better understand the impact of unexpected events on the overall trend of the data and adjust forecasting models accordingly.

The detected anomalies are then visualized in a time series plot, highlighting points of significant deviation to facilitate further investigation and model adjustments (Aggarwal, 2017).

### 3.4 Theoretical Basis of the ARIMAX model

To improve the accuracy of time series forecasting, we utilized the AutoRegressive Integrated Moving Average with eXogenous variables(ARIMAX) model, which integrates external factors into the standard ARIMA framework. This extension allows the model to account for influences beyond the inherent patterns in the target time series (Hyndman & Athanasopoulos, 2018). For this study, we explored whether including an exogenous variable, such as vaccination rates, would improve forecast accuracy compared to the ARIMA model, which relies solely on the historical values of the time series.

The ARIMAX model expands the ARIMA model by introducing exogenous regressors believed to impact the dependent variable. Mathematically, the ARIMAX model can be expressed as:

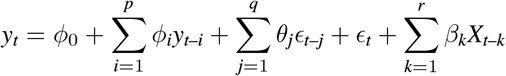

where *y*_*t*_ represents the value of the dependent variable at time *t, ϕ*_*i*_ are the autoregressive coefficients, *θ*_*j*_ are the moving average coefficients, *ϵ*_*t*_ is the error term, and *X*_*t*–*k*_ represents the exogenous variable lagged by *k* periods (Pankratz, 1991). The exogenous variables are incorporated to capture additional influences on the time series that are not explained by the series’ own historical values (Box et al., 2015).

The ARIMAX model fitting was conducted using an automated selection of ARIMA parameters (*p, d, q*), while incorporating the exogenous variable. To evaluate the performance of the ARIMAX model, we compared it with the ARIMA model using standard evaluation metrics such as Akaike Information Criterion (AIC), Root Mean Squared Error (RMSE), and Mean Absolute Error (MAE). Since these metrics were introduced earlier, they will not be repeated here (Chai & Draxler, 2014; Hyndman & Athanasopoulos, 2018).

Model forecasts were generated for a holdout period to assess prediction accuracy. The inclusion of exogenous variables in the ARIMAX model allowed us to examine whether external factors could improve the forecasting performance, providing a more comprehensive understanding of the dynamics affecting the time series. The comparison between ARIMA and ARIMAX models highlighted both the benefits and limitations of incorporating external information into the forecasting process (Pankratz, 1991).

### 3.5 Analysis of the Impact of Vaccination on New COVID-19 Cases

To analyze the relationship between vaccination rates and the number of new COVID-19 cases, several statistical methods were employed, including Granger causality testing, segmented regression, and regression discontinuity design (RDD). These methods help in understanding both the temporal relationships and potential causal effects of vaccination on the incidence of new cases (Box et al., 2015; Hyndman & Athanasopoulos, 2018).

#### 3.5.1 Granger Causality Test

To determine whether past values of the number of people vaccinated can help predict future values of new COVID-19 cases, we employed the Granger causality test. This test assesses whether one time series provides statistically significant information for forecasting another time series, suggesting a potential causal relationship (Granger, 1969). In this context, the null hypothesis of the Granger causality test is that vaccination rates do not Granger-cause new COVID-19 cases, implying that past vaccination rates do not provide additional predictive power for future case numbers when controlling for past case numbers. Detailed mathematical formulation of the model can be found in Appendix B.

#### 3.5.2 Segmented Regression Analysis and Chow Test

Segmented regression analysis was applied to quantify the impact of vaccination on the trend of new COVID-19 cases. This method estimates changes in trends before and after an intervention, such as the introduction of a vaccination program (Wagner, Soumerai, Zhang, & Ross-Degnan, 2002). The coefficients from this analysis provide estimates of both the immediate level change and the change in trend following the intervention.

To validate the results of the segmented regression, a Chow test was conducted to check for the presence of a structural break at the intervention point. The Chow test determines whether the relationship between time and new COVID-19 cases differs significantly before and after the intervention (Chow, 1960). A rejection of the null hypothesis in this test would suggest a significant change in the trend post-intervention. Detailed mathematical formulation of the segmented regression model and the Chow test can be found in Appendix B.

#### 3.5.3 Regression Discontinuity Design (RDD)

A Regression Discontinuity Design (RDD) was employed to estimate the causal effect of vaccine introduction on new COVID-19 cases, using the initiation of mass vaccination as a cutoff point (Imbens & Lemieux, 2008). RDD assumes that units on either side of the cutoff are comparable, except for the treatment. The treatment effect is captured by comparing new COVID-19 cases just before and after the introduction of the vaccination. The parameter of interest, *β*, represents the effect of the intervention at the cutoff. A non-parametric approach was used to allow for flexibility in the functional form of the relationship between time and new cases on either side of the cutoff. Details of the mathematical formulation and implementation of the RDD model can be found in Appendix B.

### 3.6 Regression Analysis of COVID-19 Infection Rates and Determinants

#### 3.6.1 Linear Regression Analysis of COVID-19 Infection Rates and Economic Development

To investigate the relationship between COVID-19 infection rates and economic development, a linear regression analysis was conducted using the infection rate as the dependent variable and GDP per capita as the independent variable. This analysis aimed to assess whether a country’s economic development is associated with its COVID-19 infection rate. Additionally, Pearson, Spearman, and Maximal Information Coefficient (MIC) were calculated to measure the strength and direction of the association between these variables (Mukaka, 2012). The Pearson and Spearman correlation coefficients assess the strength of linear and monotonic relationships, respectively, while MIC is used to detect both linear and nonlinear associations. The detailed mathematical formulation of the regression model, the correlation analyses, and MIC computation can be found in Appendix B.

#### 3.6.2 Multiple Regression Analysis with Additional Socioeconomic and Health Variables

To further explore the determinants of COVID-19 infection rates, a multiple regression model was employed, which included additional variables such as the Human Development Index (HDI), Gini coefficient, per capita health expenditure, hospital beds per 1,000 people, and population density. This model aims to assess the relative importance of various socioeconomic and health factors in explaining differences in infection rates across countries (Kutner, Nachtsheim, Neter, & Li, 2005). Interaction terms were also included to examine potential synergistic effects between variables (Montgomery, Peck, & Vining, 2012). The detailed mathematical formulation of the expanded regression model can be found in Appendix B.

#### 3.6.3 Model Selection and Multicollinearity Diagnostics

Given the potential for multicollinearity among the predictors, stepwise regression was employed to refine the model by selecting the most significant variables. Stepwise regression iteratively adds or removes predictors based on their statistical significance, optimizing the model for the lowest Akaike Information Criterion (AIC) (Burnham & Anderson, 2004).

Multicollinearity was further assessed using Variance Inflation Factor (VIF), with values greater than 10 indicating significant multicollinearity (O’Brien, 2007). To address multicollinearity, Principal Component Regression (PCR) and Partial Least Squares (PLS) regression were utilized. These methods reduce the dimensionality of the predictor space by creating uncorrelated components that explain the variance in the dependent variable (James, Witten, Hastie, & Tibshirani, 2013).

#### 3.6.4 Principal Component Regression (PCR) and Partial Least Squares (PLS) Regression

To mitigate multicollinearity and enhance the interpretability of the regression model, Principal Component Regression (PCR) and Partial Least Squares (PLS) regression were employed. Both methods involve transforming the original predictors into a smaller set of uncorrelated components, which are then used to predict the dependent variable (Jolliffe, 2002). PCR focuses on using the principal components extracted from the predictor variables, while PLS takes into account the covariance between the predictors and the dependent variable during component extraction (Wold, Sjöström, & Eriksson, 2001). Cross-validation was performed to determine the optimal number of components, with model evaluation based on minimizing the Mean Squared Error of Prediction (MSEP). Details of the PCR and PLS regression models and their formulation can be found in Appendix B.

### 3.7 Spatial Autocorrelation and Hotspot Analysis of COVID-19 Cases

In this study, spatial analysis techniques were applied to examine the distribution of COVID-19 infection rates across different regions. The analysis involved calculating Moran’s I for global spatial autocorrelation and performing the Getis-Ord Gi* statistic to identify local hotspots and coldspots. Visualization of the results was conducted using traditional red-blue color schemes, effectively highlighting areas with significant spatial clustering of high or low infection rates (Anselin, 1995; Ord & Getis, 1995).

#### 3.7.1 Spatial Autocorrelation: Moran’s I

Moran’s I is a widely used measure of global spatial autocorrelation that quantifies the degree of spatial clustering in a variable across geographic space (Cliff & Ord, 1981). It tests whether similar values (e.g., infection rates) tend to cluster spatially. A positive Moran’s I suggests that similar values cluster together, while a negative value indicates that dissimilar values are adjacent. For this analysis, a spatial weights matrix was generated based on shared boundaries between geographic regions, and Moran’s I was computed to assess the overall spatial autocorrelation of COVID-19 infection rates (Anselin, 1995). The detailed mathematical formulation of Moran’s I can be found in Appendix B.

#### 3.7.2 Hotspot Analysis: Getis-Ord Gi* Statistic

The Getis-Ord Gi* (G-star) statistic is a local spatial statistic used to identify geographic hotspots and coldspots, representing areas with significant clustering of high or low values, such as COVID-19 infection rates. Hotspots indicate clusters of high values, while coldspots represent clusters of low values. The significance of these clusters is determined through comparison with a reference distribution under the null hypothesis of spatial randomness (Getis & Ord, 1992). For this analysis, the Getis-Ord Gi* statistic was computed using a spatial weights matrix, identifying regions with statistically significant clustering of high or low infection rates (Ord & Getis, 1995). Details of the mathematical formulation of the Getis-Ord Gi* statistic can be found in Appendix B.

## 4 RESULTS AND DISCUSSION

### 4.1 Short-Term Forecasting with ARIMA Models and Anomaly Detection

To evaluate the short-term predictive performance of ARIMA models on COVID-19 case counts, forecasts were generated for four distinct periods using training data from prior months. The predictive performance was then evaluated against the actual observed data.

The first forecast, covering September 27 to December 27, 2020, was based on data from January 5, 2020, to September 27, 2020. As shown in Figure 2a, the forecast generally follows the actual case trajectory, though some deviations appear towards the end of the period, suggesting the model’s limitations in capturing sudden changes. The ACF and PACF plots (Figures 2b and 2c) indicate some residual autocorrelation, highlighting potential areas for model improvement. The Box-Ljung test returned a p-value of 0.3746, indicating that there is no significant residual autocorrelation.

**FIGURE 2.**
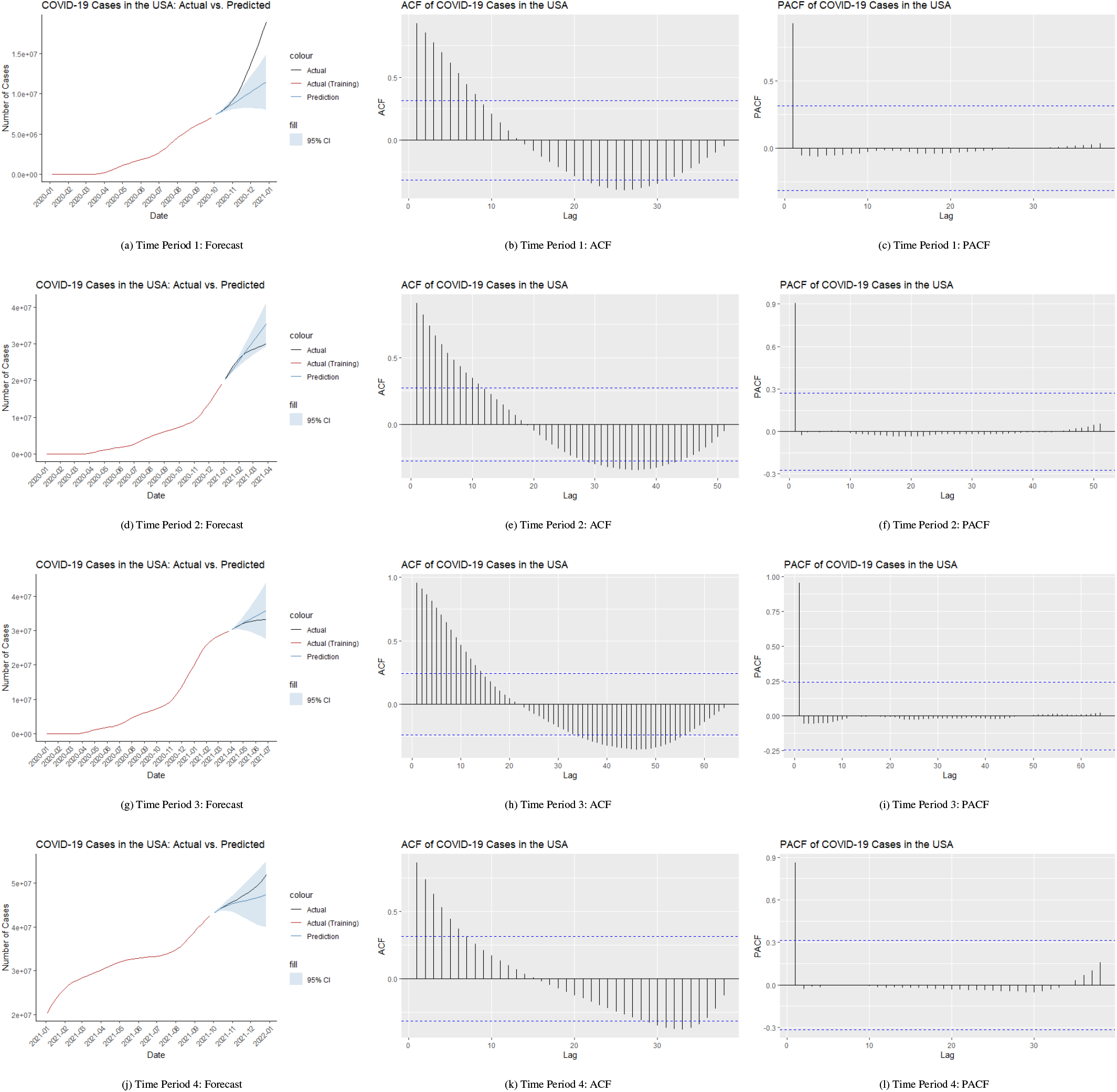
ARIMA Model Analysis in the U.S. Over Different Time Periods

For the second period, December 27, 2020, to March 28, 2021, the model was extended to include data up to December 27, 2020. Figure 2d illustrates a closer alignment between the predicted and actual cases, with only minor deviations. The ACF and PACF plots (Figures 2e and 2f) further support the model’s adequacy, though some residual correlations persisted. The Box-Ljung test for this period yielded a p-value of 0.6327, further indicating that residual autocorrelation is not a concern.

The third forecast, covering March 28 to June 27, 2021, used data up to March 28, 2021. As depicted in Figure 2g, the model closely tracks the actual case counts, demonstrating robust predictive capability. The corresponding ACF and PACF plots (Figures 2h and 2i) show that the model effectively captures the data’s temporal structure, though the Box-Ljung test resulted in a p-value of 0.07281, suggesting that there may still be some minor residual autocorrelation.

Finally, the model was applied to forecast cases from September 26 to December 26, 2021, using training data from January 3, 2021, to September 26, 2021. As seen in Figure 2j, the model continues to perform well, closely aligning with the observed case counts during the forecast period. The ACF and PACF plots (Figures 2k and 2l) indicate that the model has successfully captured the underlying patterns, with the Box-Ljung test returning a p-value of 0.2876, indicating minimal residual autocorrelation.

Throughout these periods, the ARIMA model demonstrated consistent predictive accuracy, although the residual autocorrelation evident in the ACF and PACF plots suggests areas where further refinement could enhance the model’s performance.

Among the four distinct periods analyzed using ARIMA models, the forecast for the interval from September 27 to December 27, 2020, stands out as the least accurate. Several factors may contribute to this discrepancy between the forecasted and actual data during this period.

One possible reason for the inaccuracy could be the inherent limitations of the ARIMA model, which is a linear model designed to predict future values based on past data. This model may struggle to capture sudden, nonlinear changes or external shocks that occur during the forecast period. ARIMA models assume a level of stationarity in the data, and if the underlying time series experiences structural breaks or sudden shifts, the model’s predictions can become less reliable.

Another contributing factor could be the presence of significant outliers or unexpected spikes in COVID-19 cases during the forecast period. Such anomalies might arise due to the emergence of new virus variants, changes in public health policies, or sudden shifts in public behavior, all of which can lead to rapid increases in case numbers that the model, trained on historical data, might not adequately predict.

To investigate this hypothesis, an outlier detection analysis was conducted on the data from January 5, 2020, to December 27, 2020. The detected outliers, shown in Table 2 and illustrated in Figure C2, highlight several key dates where significant anomalies were observed. These anomalies correspond to periods of sharp increases in case counts, suggesting that the forecast discrepancies could be attributed to these sudden and unexpected changes.

**TABLE 2.** COVID-19 Cases in the USA: 2020-01-05 to 2020-12-27 (Detected Outliers)

| Date Reported | Cumulative Cases |
| --- | --- |
| 2020-11-08 | 9,920,253 |
| 2020-11-15 | 10,925,098 |
| 2020-12-13 | 16,012,396 |

As shown in Table 2, significant outliers were detected on November 8, November 15, and December 13, 2020, where the cumulative cases sharply increased. These dates likely correspond to specific events or conditions that triggered a surge in cases, such as the spread of more transmissible variants or changes in testing or reporting practices.

To explore potential anomalies in COVID-19 case trends, an outlier detection analysis was performed on the cumulative COVID-19 case data for both the United States and globally. The goal was to identify points in time where the actual case numbers significantly deviated from the expected trend, potentially indicating periods when new variants emerged and spread.

The results of the outlier detection for the United States from January 5, 2020, to December 27, 2020, are shown in Figure 3a, with the detected outliers summarized in Table C1a. Notably, several of these dates align with the emergence of significant COVID-19 variants, such as the Omicron variant (B.1.1.529), first identified in November 2021 in South Africa and Botswana (Organization, 2021). Other variants, like BQ.1 and BQ.1.1, began spreading rapidly in late 2022, contributing to the increased number of cases that may have led to forecast inaccuracies (Organization, 2022a). The figure 3b shows Time Series Plot with Detected Outliers for USA.

**FIGURE 3.**
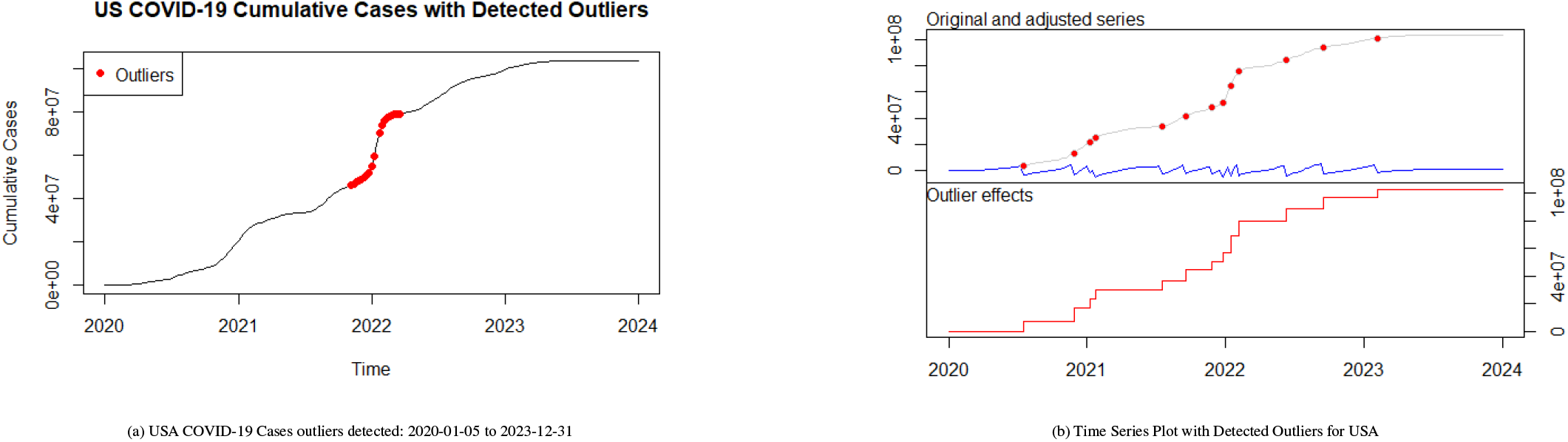
Side-by-side comparison of outliers detected and time series plot for USA

Further analysis was conducted on a global scale, with the results depicted in Figure C1. The corresponding dates and case numbers for these global outliers are listed in Table C1b. Similar to the U.S. data, these global outliers coincide with key dates when variants such as XBB, CH.1.1, and BF.7 were identified and began spreading across multiple regions, leading to significant surges in cases (Organization, 2022b). These variants, first noted in late 2022 and early 2023, had a notable impact on regions such as Asia and Europe, leading to substantial deviations from the predicted trends (Organization, 2023).

The detected outliers in both the U.S. and global datasets underscore the significant impact of COVID-19 variant emergence on the spread of the virus. Although the Alpha variant (B.1.1.7) and Gamma variant (P.1) were not explicitly captured by the outlier detection process due to their emergence towards the end of 2020, the general trend observed in Figure 3a (the U.S. outlier detection graph) does indicate a notable spike in cases during this period (Organization, 2021). This spike aligns with the timeline of Alpha and Gamma variants becoming prevalent, suggesting that the rapid spread of these variants, coupled with their increased transmissibility and potential for immune escape, likely contributed to the observed surge in case numbers. Consequently, almost all significant spikes in the data, as identified through outlier detection, correspond closely with the emergence of a new COVID-19 variant.

The dates of these outliers, as presented in Tables 1 and 2, closely match the timelines for variant identification and global spread. For example, the spikes in U.S. case counts from November 2021 to February 2022 correspond with the rise of Omicron and its subvariants (Organization, 2022a). Similarly, the global spikes identified in late 2021 and throughout 2022 align with the spread of Omicron and its subvariants, reinforcing the idea that these variants significantly impacted the accuracy of forecasted versus actual case numbers.

### 4.2 Regional COVID-19 Forecasting Across Continents

The ARIMA model was employed to forecast COVID-19 cases across different continents, including Asia, Europe, Africa, the Americas, and South America. The forecasting approach for South America involved excluding Canada, the United States, and Mexico from the Americas dataset to better approximate the COVID-19 trajectory specific to South America. The forecast results for each continent are illustrated in the figure D3, with predictions covering the period from January 2020 to early 2021.

In Asia, the ARIMA model’s predictions aligned closely with the observed data, capturing the overall upward trend in COVID-19 cases. The prediction intervals encompassed the actual case numbers, indicating the model’s robustness in this region.

In Europe, the ARIMA model’s predictions were less accurate, as the predicted cases significantly deviated from the actual observed data. This discrepancy is particularly evident towards the end of 2020, where a sharp and sudden increase in cases occurred, which the ARIMA model failed to predict effectively. Upon reviewing the forecast and considering our earlier anomaly detection for the U.S. and global data, alongside reports from the WHO regarding the emergence of variants, it is plausible to attribute this rapid rise to the Alpha variant (B.1.1.7). This variant, first detected in September 2020 in the United Kingdom, was the first to be classified by the WHO as a ”variant of concern.” The Alpha variant’s high transmissibility likely contributed to the significant increase in cases, which was not fully captured by the ARIMA model, highlighting the challenges of forecasting during periods of rapid epidemiological change.

For Africa, the model demonstrated a good fit with the actual data, though, like in Europe, the rapid rise in cases near the year’s end pushed the limits of the prediction interval. In the Americas, the ARIMA model performed strongly, with predictions closely matching the steep increase in case numbers. This region, which experienced one of the most significant surges in cases, was effectively modeled, with predictions falling within the confidence intervals.

In South America, after excluding the northern countries, the ARIMA model continued to perform well. The predicted cases remained within reasonable bounds compared to the observed data, similar to the other continents.

Across all regions, the Box-Ljung test p-values were significantly above the 0.05 threshold, indicating no significant auto-correlation in the residuals. This suggests that the ARIMA models successfully captured the temporal patterns of COVID-19 case progression in each region. The occasional underestimations, particularly during periods of rapid increases in cases, underscore the challenges posed by the pandemic’s dynamic nature and the emergence of new variants, which may not have been fully accounted for in models trained on earlier data. Nonetheless, the overall performance of the ARIMA models across these diverse regions was robust, providing valuable insights into the spread of COVID-19 during the forecasted periods.

### 4.3 Rolling Window Cross-Validation and Comparison with auto.arima

In our previous ARIMA forecasting efforts, we relied on the `auto.arima` function to automatically select the ARIMA model parameters *p, d*, and *q*. The `auto.arima` function is based on minimizing the Akaike Information Criterion (AIC), which balances model fit and complexity by penalizing excessive parameters. This approach offers several advantages, including speed, automation, and often, reasonably good results. However, its reliance on AIC may not always yield the most accurate forecasts, especially when dealing with nonstationary time series or when the model is used for long-term predictions.

To explore whether other parameter selection methods could improve forecast accuracy, we applied a rolling window cross-validation technique to optimize the *p* and *q* parameters, while keeping *d* fixed as determined by `auto.arima`. The underlying reason for fixing *d* lies in its role in differencing, which is generally determined by the data’s trend and seasonality structure-a topic well-supported by statistical theory. For example, once the time series is made stationary through differencing, the differencing order *d* should remain constant to maintain this stationarity, regardless of changes in *p* and *q*.

In our analysis, we selected a period where the ARIMA predictions notably diverged from the actual data, such as the case with COVID-19 cases in the USA and Europe from January 5, 2020, to December 27, 2020. The divergence was primarily due to the unforeseen surge in cases caused by the emergence of new variants, highlighting the limitations of traditional ARIMA models in capturing such abrupt changes.

Using the rolling window cross-validation approach, we evaluated different combinations of *p* and *q* based on the Root Mean Squared Error (RMSE) metric. This method, which assesses the out-of-sample performance of models across multiple training windows, is particularly valuable when forecasting non-stationary time series with varying patterns over time. Table 3 summarizes the RMSE values for the USA’s ARIMA model with parameters selected via cross-validation versus those obtained using `auto.arima`, while Figure 4a provides a heatmap visualizing the RMSE across different *p* and *q* combinations.

**TABLE 3.** RMSE Comparison between auto.arima and Cross-Validation for ARIMA Models (USA).

| Model | ARIMA Parameters |  |  | RMSE |
| --- | --- | --- | --- | --- |
| | $p$ | $d$ | $q$ | RMSE |
| `auto.arima` | 1 | 2 | 0 | 27648.12 |
| Cross-Validation | 2 | 2 | 2 | 22949.3 |

**FIGURE 4.**
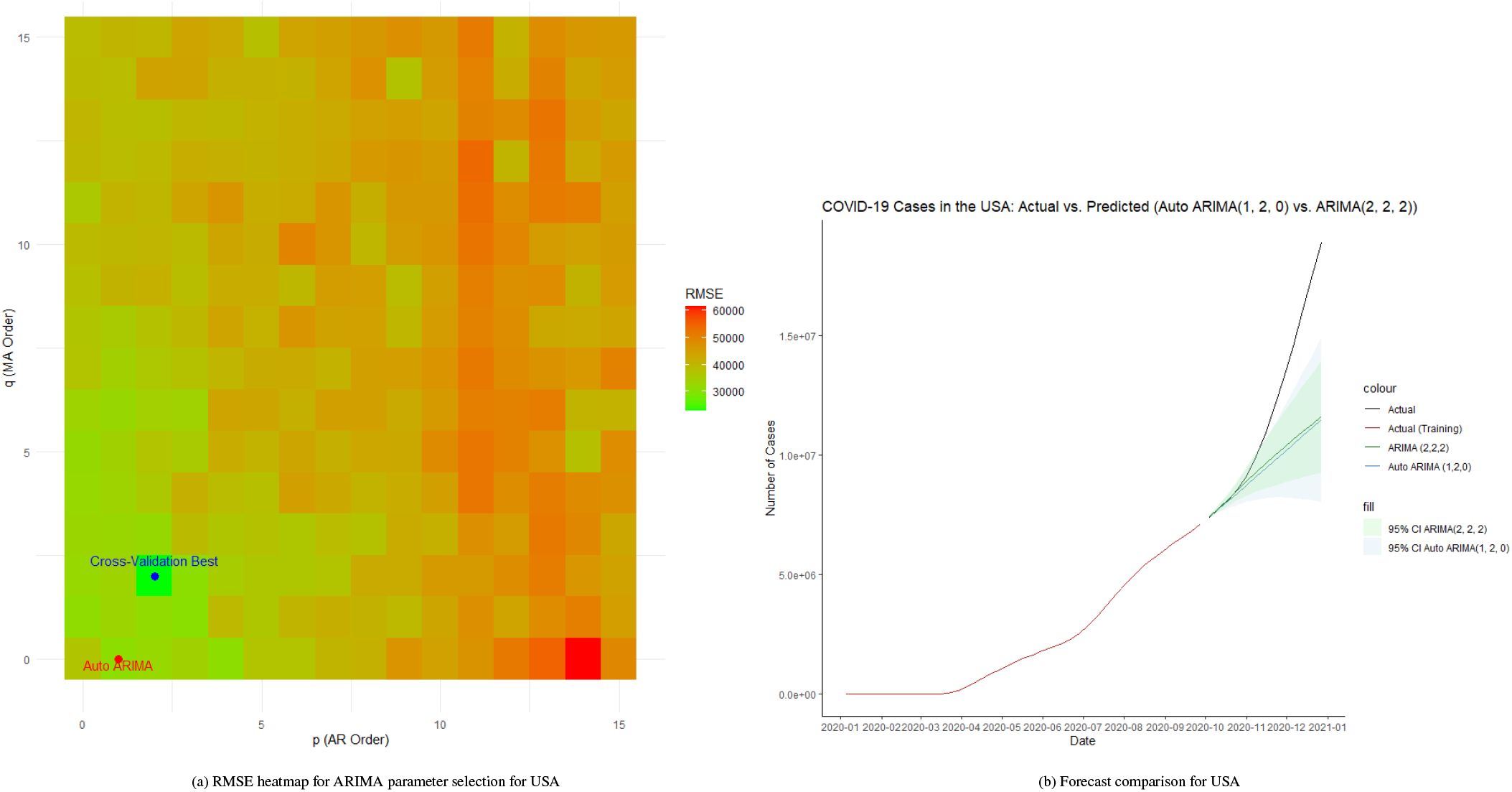
Heatmap of RMSE and forecast comparison of USA Note: The blue points represent the observed data, and the red line represents the fitted regression discontinuity model. The vertical black line indicates the intervention date (December 13, 2020).

As depicted in Figure 4a, the RMSE heatmap clearly shows that the cross-validated ARIMA parameters (*p* = 2, *q* = 2) outperform the `auto.arima` parameters (*p* = 1, *q* = 0) in terms of RMSE. The heatmap provides a comprehensive view of how different combinations of *p* and *q* impact the forecast accuracy, with lower RMSE values indicating better performance.

Furthermore, Figure 4b compares the forecasted COVID-19 cases in the USA using the `auto.arima` model and the cross-validated ARIMA model. Although both models exhibit significant deviations from the actual data due to the unexpected surge in cases, the cross-validated model’s predictions are slightly closer to the actual values than those of `auto.arima`, suggesting that the cross-validation approach can yield more accurate forecasts under certain conditions.

For the ARIMA model applied to Europe, a similar approach was employed as described in the previous. Table E2 presents the RMSE values for the European ARIMA model, comparing the parameters selected via cross-validation with those determined by `auto.arima`. The RMSE heatmap shown in Figure E4a visualizes the performance across different combinations of *p* and *q*. Figure E4b illustrates the comparison of the forecasted COVID-19 cases in Europe using the ARIMA model with parameters selected by cross-validation against those obtained by `auto.arima`. As observed, the forecast line derived from the cross-validated model is significantly closer to the actual data than the one produced by `auto.arima`, although both models still show notable deviations from the actual trajectory. These findings closely mirror the conclusions drawn from the cross-validation results for the ARIMA model applied to the USA, further reinforcing the potential advantages of cross-validation in parameter selection for ARIMA models in the context of highly volatile and non-stationary time series data.

### 4.4 Effect of Vaccination on New COVID-19 Cases

From December 2020, the global effort to vaccinate against COVID-19 began, raising the critical question of whether the vaccination campaign effectively reduced the number of new COVID-19 cases. To address this question, several statistical methods were employed, including the Granger Causality Test, segmented regression analysis, the Chow Test, and Regression Discontinuity Design (RDD).

The analysis began with the Granger causality test, aimed at determining whether the number of people vaccinated could serve as a predictor for future new COVID-19 cases, while controlling for past cases. Two models were compared: the first model included lags of both new cases and vaccinations, while the second model included only lags of new cases. The results did not indicate a significant causal relationship between vaccination and a reduction in new cases, as the F-statistic was 0.24 with a p-value of 0.9746, suggesting that the inclusion of vaccination data did not improve the predictive power of the model within the lags tested. Specifically, with a lag of 7 (equivalent to 49 days), the Granger causality test showed no significant effect of vaccination on new cases within this period. As shown in table4.

**TABLE 4.** Granger Causality Test Results.

| Model | Res.Df | Df | F | Pr(>F) |
| --- | --- | --- | --- | --- |
| Model 1: `new_cases ~ Lags(new_cases, 1:7) + Lags(people_vaccinated, 1:7)` | 151 |  |  |  |
| Model 2: `new_cases ~ Lags(new_cases, 1:7)` | 158 | -7 | 0.24 | 0.9746 |

To further investigate the potential impact of vaccination on the trend in COVID-19 cases, segmented regression analysis was employed, introducing a breakpoint at the onset of the vaccination campaign. The regression model accounted for time, an indicator for the postintervention period, and the interaction between time and the post-intervention phase. The analysis revealed that while the overall trend in new cases showed a positive slope (*β* = 18987, p = 0.02136), the interaction term (`time_post_intervention`) was negative and significant (*β* = –24115, *p* = 0.00445), indicating a deceleration in the growth rate of new cases following the intervention. Despite this, the post-intervention indicator itself was not statistically significant (*p* = 0.31000), which aligns with the results of the Granger causality test, further suggesting that the immediate effect of vaccination on reducing new cases was not significant. However, the negative and significant interaction term implies that vaccination had a significant long-term impact on reducing new cases, indicating a beneficial effect over time. Figure 5a illustrates the segmented regression results, showing how the predicted number of cases diverges from the actual cases over time. As shown in Table F3, the segmented regression results demonstrate these trends clearly.

**FIGURE 5.**
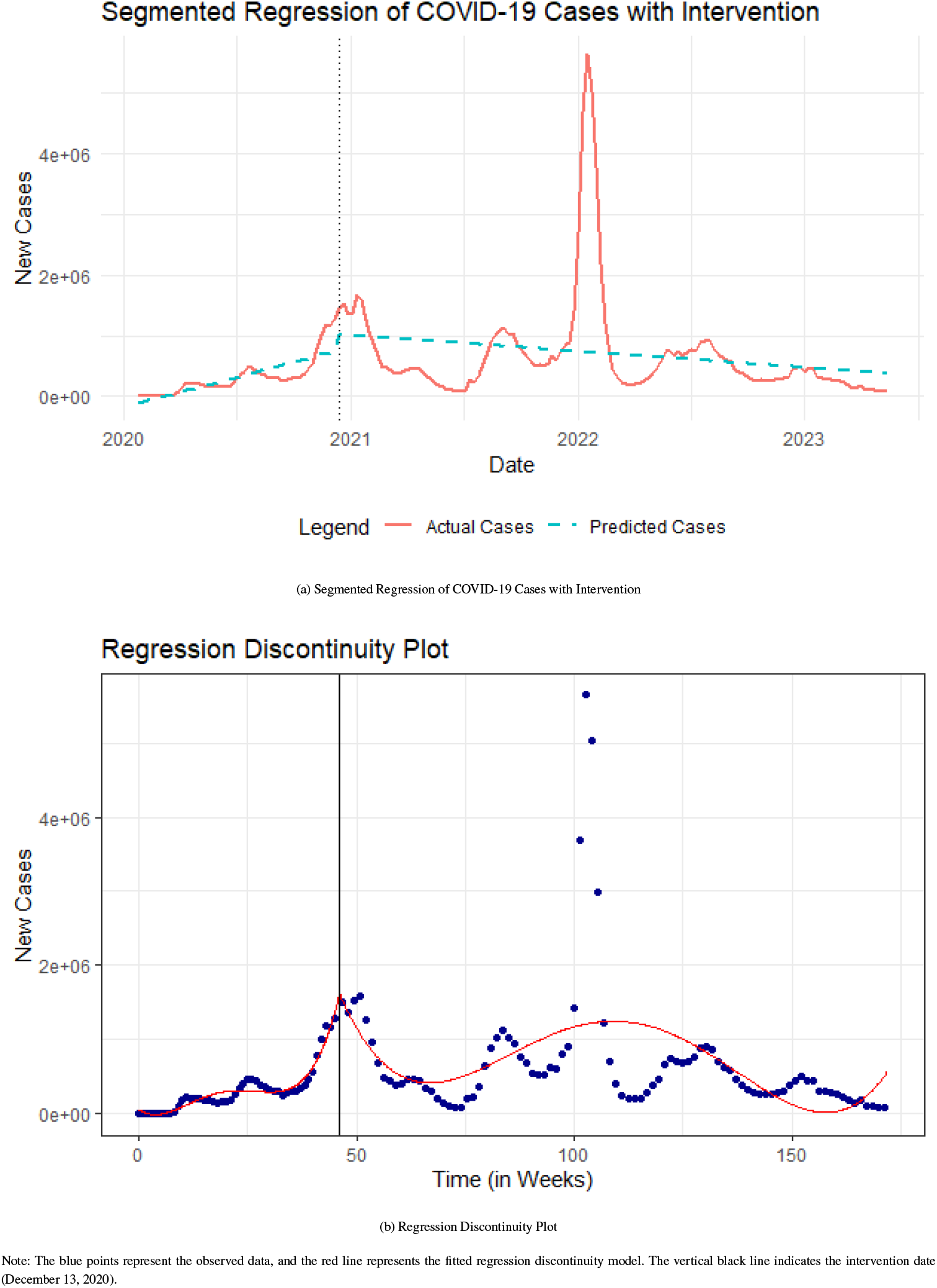
Comparison of Segmented Regression of COVID-19 Cases and Regression Discontinuity Plot

Additionally, the Chow Test was conducted to formally assess the presence of a structural break at the intervention point. The test provided strong evidence of a structural change, with a p-value of 6.437e-06, indicating that the introduction of the vaccination campaign significantly altered the underlying relationship between time and new cases. This result corroborates the findings from the segmented regression analysis, suggesting that vaccination led to a structural shift in the trend of new cases. Finally, a Regression Discontinuity Design (RDD) was applied to further validate these findings. The RDD analysis, focusing on the sharp change in the trend of new cases at the point of intervention, yielded a conventional coefficient estimate of 76662.154 with a non-significant p-value (p = 0.636). This suggests that while there may have been an observable shift in the trend of new cases at the intervention point, it was not statistically significant at conventional levels. As shown in Table F4, the regression discontinuity results further support the conclusion that the immediate impact of vaccination was not statistically significant. Figure 5b provides a visualization of the RDD results, highlighting the discontinuity at the intervention point. The non-significant result from the RDD analysis is consistent with the findings from both the Granger causality test and the segmented regression, indicating that the immediate impact of vaccination was not significant.

### 4.5 Forecast on COVID-19 cases using ARIMAX with vaccine as an exogenous variable

Building upon the results in the previous section, where it was demonstrated that vaccination had a significant long-term impact on the reduction of new COVID-19 cases, a logical extension was to incorporate the number of vaccinations as an exogenous variable in ARIMAX models. The hypothesis was that the inclusion of this variable might improve the accuracy of forecasts compared to a standard ARIMA model, which does not account for such external factors.

To test this hypothesis, we utilized our dataset where vaccination began on December 13, 2020. Given the evidence that the impact of vaccination is more pronounced over the long term, the first training period selected for the model was from January 5, 2020, to June 27, 2021, approximately six months after the start of vaccination. This period was used to predict the cumulative number of cases for the subsequent three months. Subsequently, the training data span was gradually extended in two further scenarios:

1. January 5, 2020 – June 27, 2021
2. January 5, 2020 – December 26, 2021
3. January 5, 2020 – September 25, 2022

Figures 6, G5, and G6 respectively compare the ARIMA and ARIMAX model predictions for these three periods, and tables 5, G5, G6 compare the metrics of ARIMA and ARIMAX models across three time periods, including AIC, RMSE, and MAE.

**TABLE 5.**
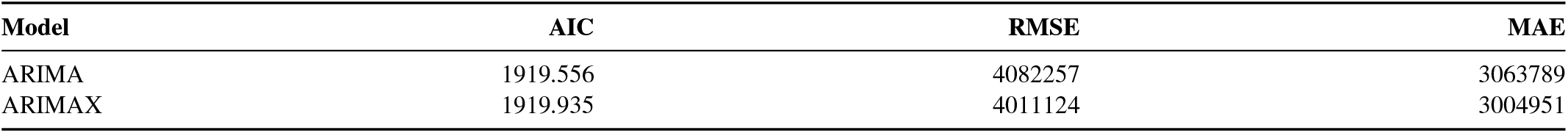
Comparison of ARIMA and ARIMAX Models (Period 1).

**FIGURE 6.**
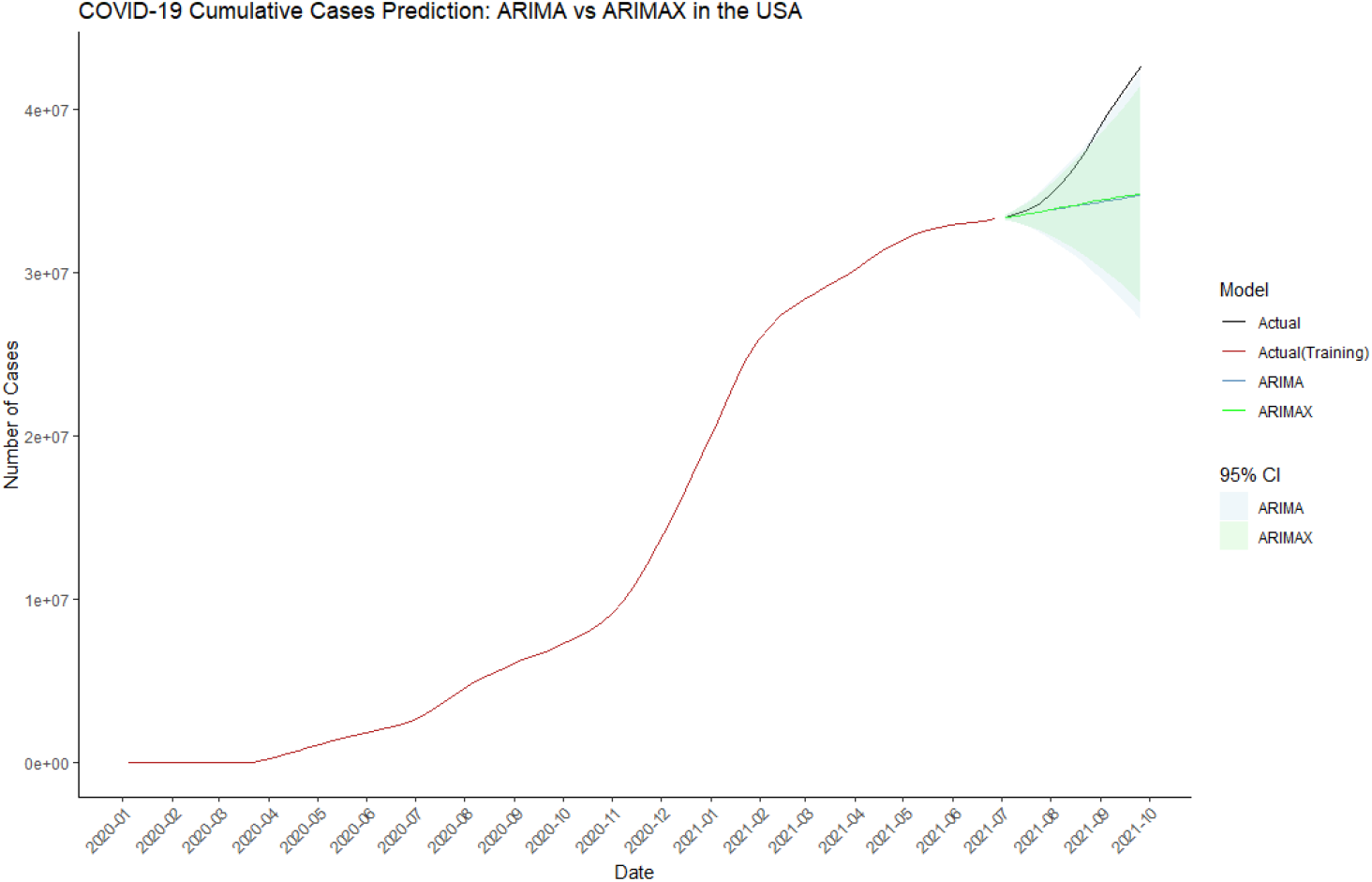
ARIMAX forecast for period 1

From these results, it can be observed that the ARIMAX model sometimes produces forecasts closer to the actual data than the ARIMA model, as indicated by lower RMSE and MAE values in certain scenarios. However, there are also cases where the ARIMAX model deviates more from the actual data, resulting in higher RMSE values. Interestingly, the Akaike Information Criterion (AIC) does not always correlate directly with RMSE and MAE improvements. For example, in the third period, despite the ARIMAX model providing predictions closer to the actual data (as reflected by lower RMSE and MAE), its AIC is higher than that of the ARIMA model. This higher AIC reflects the trade-off between model complexity and goodness-of-fit inherent in the AIC calculation.

### 4.6 Multivariate Regression Analysis of Global COVID-19 Infection Rates

To investigate the factors influencing COVID-19 infection rates across different countries, we began by hypothesizing that more developed countries, with their advanced medical infrastructure and higher availability of healthcare resources, might exhibit lower infection rates. However, an examination of the top 10 countries by infection rate as of December 31, 2023 (Figure H7), revealed that many of the countries with the highest infection rates are, in fact, highly developed. For example, countries such as Luxembourg, Denmark, and Austria, all of which are considered highly developed, show among the highest infection rates, challenging the initial hypothesis.

To explore this relationship further, we conducted a linear regression analysis between the COVID-19 infection rate and GDP per capita as a proxy for a country’s level of development. The scatterplot with the regression line is displayed in Figure 7. The regression output showed a significant positive relationship between GDP per capita and infection rate, with the coefficient for GDP per capita being positive and highly significant (p < 2e-16), indicating that higher GDP per capita is associated with higher infection rates. The regression model, however, had a relatively low R-squared value of 0.4763, suggesting that while GDP per capita is a significant predictor, it explains only about 47.63% of the variance in infection rates.

**FIGURE 7.**
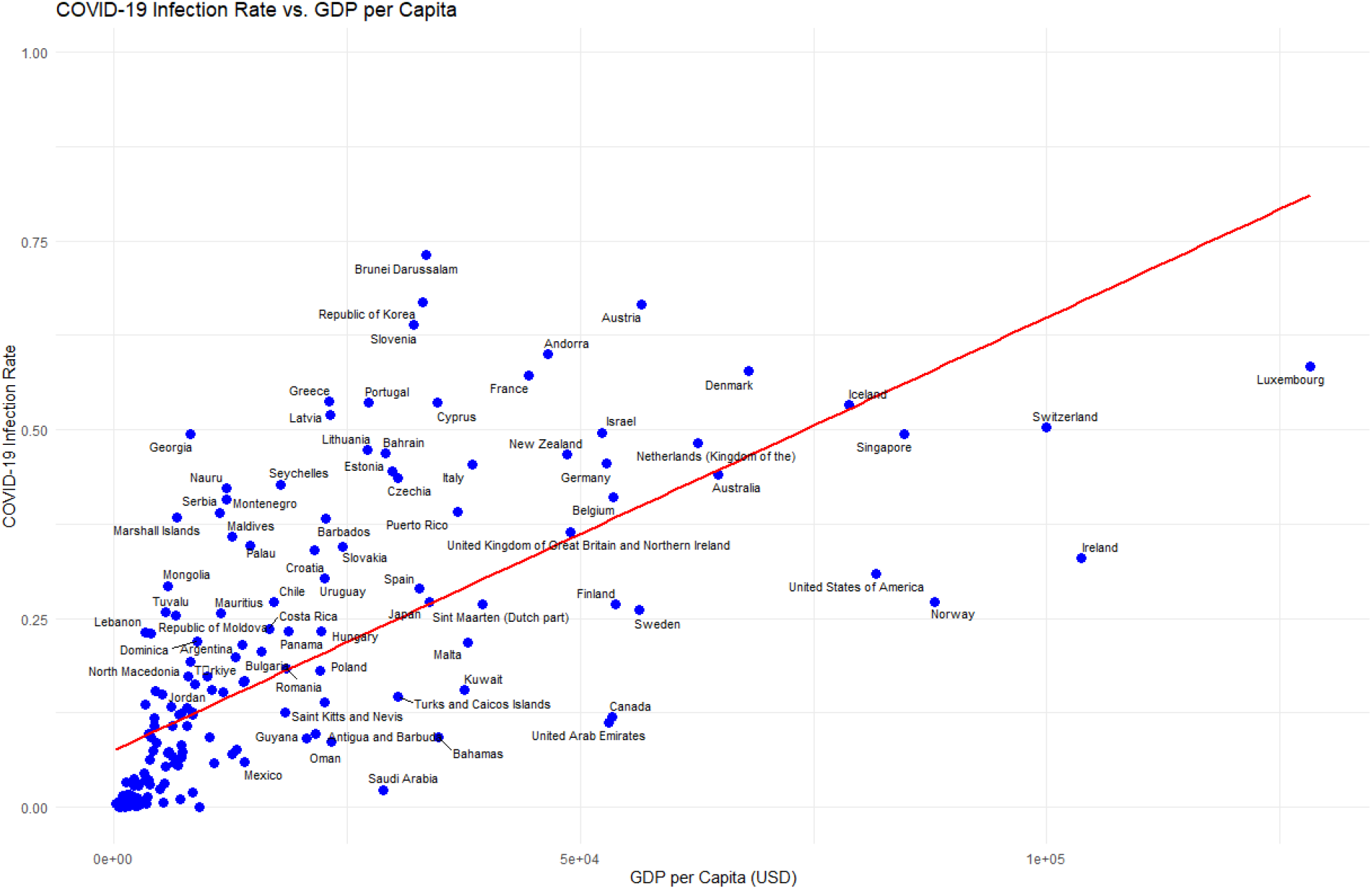
The scatterplot with regression line

In addition to the regression analysis, we calculated three correlation metrics to further understand the relationship between GDP per capita and infection rate. The Pearson correlation coefficient was 0.690161, indicating a moderately strong positive linear relationship between the two variables. The Spearman rank correlation coefficient was higher, at 0.8593242, suggesting a strong monotonic relationship. Finally, the Maximal Information Coefficient (MIC) was 0.7256325, pointing to a strong association that may capture nonlinear relationships between GDP per capita and infection rate. These correlations further support the conclusion that higher GDP per capita is associated with increased infection rates, although other factors likely contribute to the remaining unexplained variance.

Given the relatively low R-squared, it was clear that other factors beyond GDP per capita might influence infection rates. Therefore, we expanded the model to include additional variables that could plausibly affect infection rates: Human Development Index (HDI), Gini coefficient (a measure of income inequality), per capita health expenditure, the number of hospital beds per 1,000 people, and population density. The resulting multivariate regression model incorporated both main effects and interaction terms between these variables.

The multivariate regression results, which included interaction terms, revealed a more complex relationship between the predictors and the infection rate. While GDP per capita continued to have a significant effect (p = 0.006544), other variables like health expenditure and certain interaction terms also emerged as significant predictors. For example, the interaction between GDP per capita and HDI (p = 0.006381) and between GDP per capita and the Gini coefficient (p = 0.029676) were both significant, indicating that the effect of GDP per capita on infection rates is moderated by a country’s HDI and income inequality. Additionally, the interaction between HDI and health expenditure (p = 0.000724) was significant, suggesting that the combined effect of these two factors significantly influences infection rates. The detailed results of the regression analysis, including estimates, standard errors, t-values, and p-values, are provided in Table I7.

Despite these findings, the model’s R-squared increased substantially to 0.8179, indicating that approximately 81.79% of the variance in infection rates can be explained by the expanded set of predictors and their interactions. However, the residual plots (Figure 8a) reveal some potential issues with model fit, including non-constant variance (heteroscedasticity) and some deviation from normality in the residuals, as indicated by the Q-Q plot.

**FIGURE 8.**
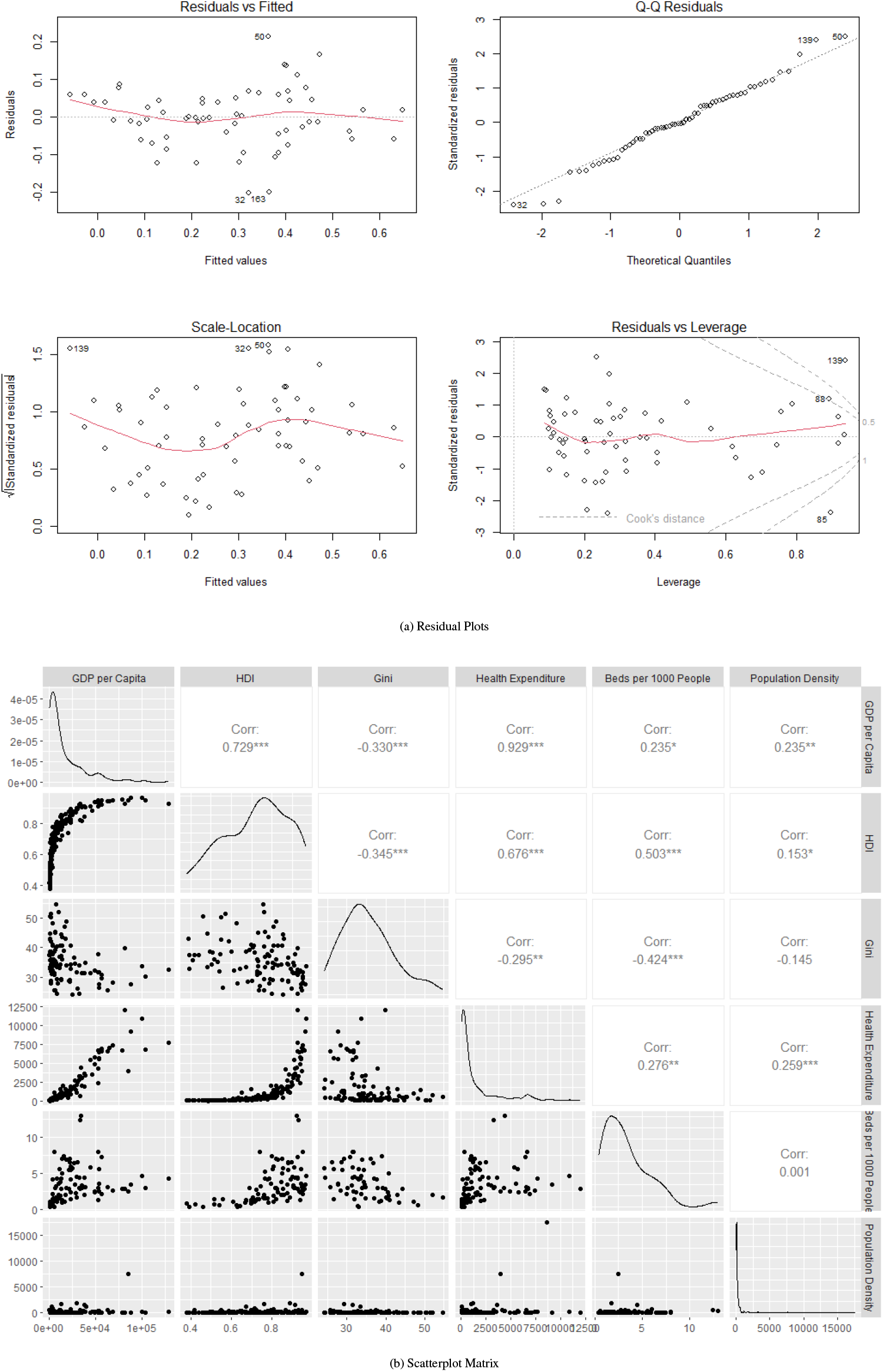
Comparison of Residual Plots and Scatterplot Matrix

The scatterplot matrix (Figure 8b) and coefficient plot (Figure J8) further underscore the complexity of the relationships among the predictors. The scatterplot matrix highlights the correlations between variables, with some expected relationships, such as the positive correlation between GDP per capita and HDI (corr = 0.729) and a negative correlation between GDP per capita and the Gini coefficient (corr = -0.330). The coefficient plot shows the magnitude and direction of the effects, with GDP per capita, health expenditure, and certain interaction terms having the most pronounced impacts on infection rates.

#### 4.6.1 Addressing Multicollinearity in the Regression Model

The initial multivariate regression model that included interaction terms significantly improved the model’s explanatory power, as indicated by a substantial increase in the *R*^2^ value. However, this complexity introduced severe multicollinearity into the model, as evidenced by the extremely high Variance Inflation Factor (VIF) values. For instance, variables such as GDP per capita, HDI, and health expenditure, along with their interaction terms, exhibited VIF values that were in the tens of thousands, indicating that multicollinearity was indeed a significant problem. Multicollinearity can render the regression coefficients unstable and their interpretation challenging, which necessitated a more rigorous approach to model simplification and stabilization.

To mitigate these issues, we employed stepwise regression as an initial step to reduce model complexity by removing less significant predictors. The simplified model retained key variables and interactions while excluding those that contributed less to the model’s overall explanatory power. The resulting model, while more manageable, continued to exhibit notable multicollinearity, with several VIF values remaining high, albeit reduced from their initial levels.

Given the persistence of multicollinearity, we explored alternative methods to further address this issue. Both Partial Least Squares (PLS) and Principal Component Regression (PCR) were considered, as these techniques are specifically designed to handle multicollinearity by transforming the predictor variables into a set of uncorrelated components.

We applied PLS and PCR to the dataset, each method aiming to reduce the dimensionality of the predictor variables while maximizing the explained variance in the response variable, infection rate. The PLS analysis, as shown in Table 7, was particularly effective in this context. The model explained 67.22% of the variance in infection rates using five components, which was identified as the optimal number of components through cross-validation (Figure 9). Beyond five components, the model’s Mean Squared Error of Prediction (MSEP) began to increase, suggesting that additional components might introduce noise rather than improve predictive accuracy.

**TABLE 6.**
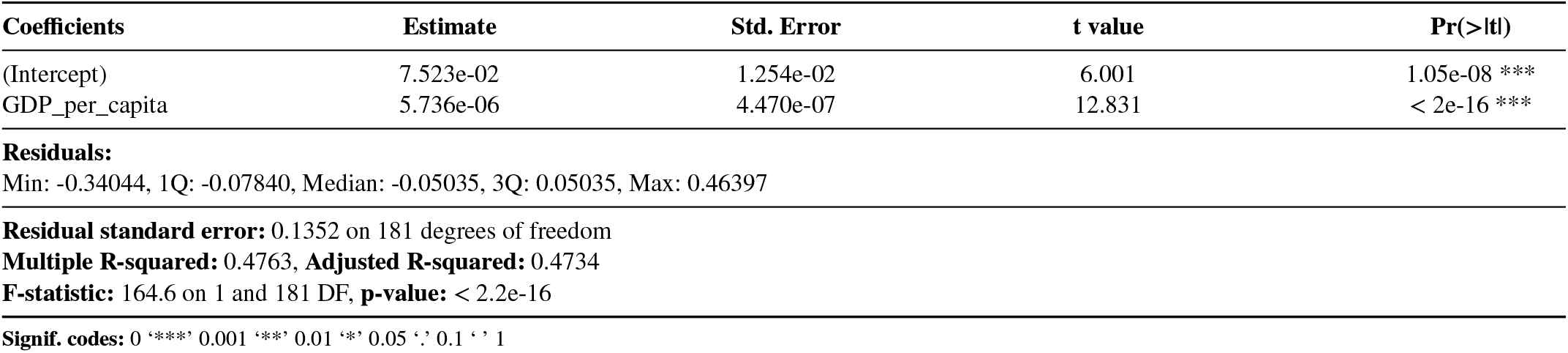
Linear Regression Results: Infection Rate vs. GDP per Capita.

**TABLE 7.** PLS Results: Variance Explained by Number of Components.

|  | 1 comps | 2 comps | 3 comps | 4 comps | 5 comps | 6 comps |
| --- | --- | --- | --- | --- | --- | --- |
| X | 47.98 | 66.98 | 81.77 | 88.76 | 94.81 | 97.28 |
| infection_rate | 48.67 | 55.79 | 59.75 | 65.34 | 67.22 | 70.14 |
|  | 7 comps | 8 comps | 9 comps | 10 comps | 11 comps |  |
| X | 98.05 | 98.92 | 99.48 | 99.78 | 99.85 |  |
| infection_rate | 72.68 | 73.76 | 73.93 | 74.10 | 74.56 |  |
|  | 12 comps | 13 comps | 14 comps | 15 comps | 16 comps |  |
| X | 99.90 | 99.95 | 99.97 | 99.98 | 99.99 |  |
| infection_rate | 75.47 | 75.98 | 76.27 | 76.49 | 76.84 |  |
|  | 17 comps | 18 comps | 19 comps | 20 comps | 21 comps |  |
| X | 99.99 | 100.00 | 100.00 | 100.00 | 100.00 |  |
| infection_rate | 77.45 | 77.79 | 78.81 | 81.62 | 81.79 |  |

**FIGURE 9.**
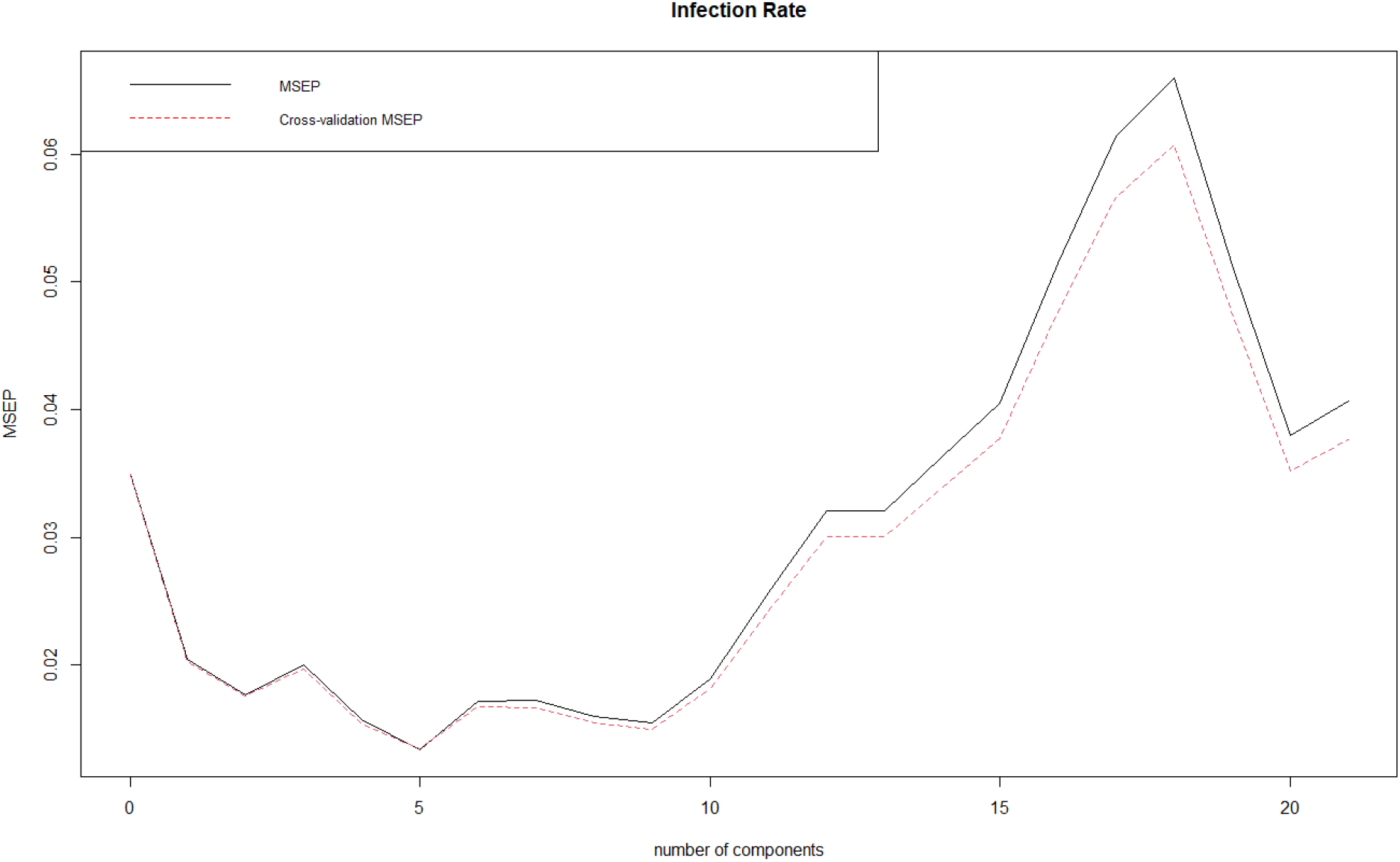
Cross-Validation Mean Squared Error Plot of the PLS model

In addition to the error plot, Table K9 provides detailed cross-validation results for each model. This table presents the MSEP for different numbers of components, highlighting how the error decreases as the number of components increases up to five, and then rises with the inclusion of additional components. These results reinforce the findings illustrated in Figure 9, where five components were found to be optimal.

The component loadings from the PLS model, visualized in the heatmap (Figure M9), highlight the contribution of each variable to the principal components. Variables such as GDP per capita, HDI, and health expenditure had substantial loadings on the first few components, indicating their significant influence on the model. However, complex interactions, such as those between GDP per capita and population density or health expenditure and population density, also played critical roles in later components.

In addition to the heatmap, the detailed PLS loadings are presented in Table L10. This table provides specific loading values for each variable across the first five components, further illustrating the contributions and interactions among variables in shaping the principal components.

The PLS model, through component reduction, provided a more stable set of coefficients, as shown by the reduced VIF values and improved interpretability of the regression coefficients. The final regression coefficients derived from the PLS model (Table L11) illustrate the direct and interaction effects of the predictor variables on infection rates, offering a clearer understanding of the underlying relationships.

PCR, as shown in Table K8, on the other hand, showed similar results but with slightly lower explained variance for the same number of components. While PCR effectively reduced multicollinearity, the trade-off was in the form of lower predictive power compared to PLS. Specifically, PCR explained 59.43% of the variance in infection rates with six components, which increased to 75.81% with 17 components, but did not surpass the PLS model in terms of overall performance.

### 4.7 Spatial Autocorrelation and Hotspot Analysis of COVID-19 Cases in U.S. States

We performed a spatial autocorrelation and hotspot analysis of COVID-19 cases in U.S. states. Spatial autocorrelation measures the degree to which COVID-19 cases are spatially clustered, while hotspot analysis identifies regions with significantly high or low case counts.

Using the most recent COVID-19 case data across U.S. states, we calculated the number of COVID-19 cases per 100,000 people to normalize for population differences. Figure 10a illustrates the spatial distribution of these normalized case counts across the United States. States with higher case rates per 100,000 people are represented in shades of red, while those with lower rates are in shades of blue. Notably, Alaska and several states in the South exhibit particularly high case rates.

**FIGURE 10.**
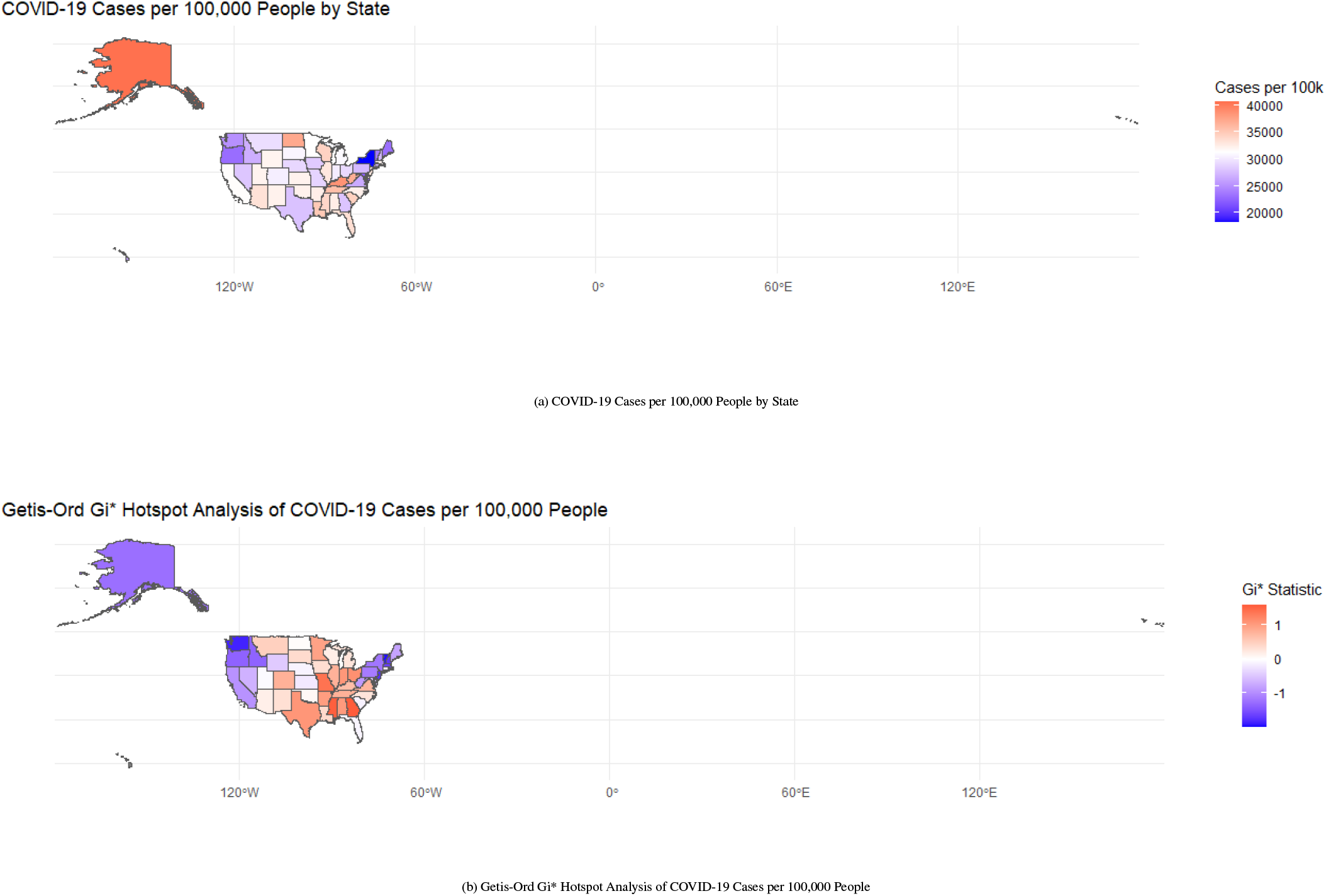
(a) COVID-19 Cases per 100,000 People by State, (b) Getis-Ord Gi* Hotspot Analysis of COVID-19 Cases per 100,000 People

In addition to the spatial visualization, Table N12 provides the detailed rankings of the top 10 and bottom 10 states in terms of COVID-19 cases per 100,000 people. Alaska ranks highest with 40,576.16 cases per 100,000 people, followed by Rhode Island and Kentucky. Conversely, New York, Maryland, and Oregon report the lowest case rates, with New York at the bottom with 18,251.51 cases per 100,000 people.

To further understand the spatial pattern, we conducted a Moran’s I test, a measure of spatial autocorrelation. The results revealed a Moran’s I statistic of 0.1578 with a p-value of 0.03169, indicating a significant positive spatial autocorrelation. This suggests that states with high COVID-19 case rates tend to be geographically clustered rather than randomly distributed.

To identify specific clusters of high or low case rates, we applied the Getis-Ord Gi* statistic, which provides a measure of local spatial clustering. As shown in Figure 10b, the hotspot analysis identified several ”hot” and ”cold” spots. Southern states such as Arkansas, Georgia, and Mississippi, as well as parts of Texas and Ohio, were identified as hotspots (with high Gi* values), indicating significant clustering of high case rates. Conversely, states like Alaska, Delaware, New Hampshire, and Vermont were identified as cold spots, suggesting significant clustering of low case rates.

In addition, Table N13 lists the top 10 hotspot and coldspot states along with their corresponding Gi* values. For instance, Arkansas has the highest Gi* value of 1.000414, marking it as a significant hotspot, while Alaska has the lowest Gi* value of -1.237323, making it a prominent coldspot. These detailed Gi* values provide a quantitative basis for understanding the spatial clustering patterns observed in the map.

For Alaska, given that it does not share borders with any other state, we designated Washington as its sole neighbor. Despite this adjustment, Alaska, which has the highest infection rate among all states, was classified as a cold spot in the Getis-Ord Gi* Hotspot analysis. This counterintuitive result may be due to the isolation of Alaska, where the lack of adjacent states diminishes the influence of its high case rate on surrounding areas. Consequently, its high infection rate is not part of a broader regional trend, leading the Gi* statistic to categorize it as a cold spot rather than a hotspot. This highlights the importance of considering geographic and relational context in spatial analyses, particularly for isolated regions.

## 5 CONCLUSIONS AND DISCUSSION

The comprehensive statistical analysis of COVID-19 trends, employing ARIMA, ARIMAX, multiple regression, and spatial autocorrelation models, provides valuable insights into the dynamics of the pandemic both globally and within the United States. These findings underscore the strengths and limitations of different modeling approaches and highlight the complexity of factors influencing COVID-19 case numbers.

The ARIMA models demonstrated robust performance in predicting short-term COVID-19 trends, particularly in scenarios where case dynamics followed relatively stable patterns (Hyndman & Athanasopoulos, 2018). However, the models exhibited limitations when faced with abrupt shifts in infection rates, such as those caused by sudden policy changes or the emergence of new virus variants (Chowell, Hyman, & Castillo-Chavez, 2021). These scenarios often resulted in less accurate forecasts, suggesting that while ARIMA models are useful for capturing general trends, they may require augmentation or combination with other models to better account for sudden, non-linear changes (Liu, Magal, Seydi, & Webb, 2020).

The ARIMAX models, which incorporate exogenous variables such as vaccination data, provided a more nuanced analysis by attempting to account for external influences on COVID-19 case numbers (Pankratz, 1991). However, the effectiveness of the ARIMAX model is highly contingent on the specific characteristics of the time period and the data involved. For instance, during periods where the impact of vaccination on case numbers is either delayed or not as pronounced, the model struggled to accurately reflect the true relationship between the variables (Li et al., 2021). This is particularly evident when vaccine uptake is gradual, or when the effects of vaccination take time to manifest in the population. Under such conditions, the model may either overestimate or underestimate the influence of vaccination, leading to skewed forecasts (Hernández-Orallo, Calafate, Cano, & Manzoni, 2022).

Several challenges were identified in the application of the ARIMAX model. Firstly, the model assumes a direct and linear impact of the exogenous variable (vaccination) on the dependent variable (COVID-19 cases), which may not fully capture the complex, non-linear relationships at play (Hyndman & Athanasopoulos, 2018). Factors such as varying vaccine efficacy, the emergence of new virus variants, changes in public behavior, and policy interventions (e.g., lockdowns, mask mandates) can all influence the effectiveness of vaccination efforts in reducing case numbers (Gao, Yang, Wang, Li, & Zhao, 2022). If these factors are not adequately incorporated into the model, the ARIMAX model might misattribute changes in case numbers to vaccination, leading to inaccurate predictions.

Moreover, the inclusion of vaccination data as an exogenous variable introduces the risk of multicollinearity, particularly if the vaccination data is correlated with other variables influencing the spread of COVID-19. Multicollinearity can lead to instability in the coefficient estimates, making the model’s predictions less reliable (O’Brien, 2007). In some cases, this instability may cause the ARIMAX model to perform worse than the simpler ARIMA model, which does not encounter this complication.

Timing also plays a crucial role in the performance of the ARIMAX model. The effects of vaccination on COVID-19 cases may exhibit variable lags that are not constant or predictable (Li et al., 2021). If the model fails to capture the appropriate lag structure, it could lead to inaccurate predictions. For example, the time required for immunity to build following vaccination or differences in how various population segments respond to vaccination can lead to mismatches between vaccination data and changes in case numbers, further complicating the accuracy of ARIMAX predictions.

Additionally, there is a risk of overfitting with the ARIMAX model, especially when the model becomes overly complex relative to the amount of available data. Overfitting occurs when the model captures noise or random fluctuations in the training data as meaningful patterns, leading to less accurate predictions when applied to new data (James et al., 2013). This is particularly problematic when incorporating vaccination data, as the added complexity could diminish the model’s generalizability.

In the multiple regression analysis, several socioeconomic factors were identified as significant predictors of COVID-19 case numbers. For example, factors such as population density, median income, and access to healthcare services showed strong correlations with case numbers (Wooldridge, 2016). These findings highlight the unequal impact of the pandemic across different demographic groups and regions. Specifically, areas with higher population density and lower income levels tended to experience higher case numbers, likely due to the increased difficulty in implementing social distancing and the limited access to healthcare services (Bambra et al., 2020).

The regression analysis further emphasized the importance of considering a broad range of socioeconomic factors when assessing the spread of COVID-19. However, the model also revealed some limitations. The relationships between the independent variables and COVID-19 case numbers were not always linear, suggesting the need for more sophisticated modeling approaches that can capture these complexities (Montgomery et al., 2012). Moreover, the presence of interaction effects among the variables, such as the combined impact of income and healthcare access, indicates that future models should explore these interactions to better understand the pandemic’s dynamics.

Spatial autocorrelation analyses provided additional insights, particularly regarding the geographic clustering of COVID-19 cases. The results highlighted significant spatial clusters of high infection rates, suggesting that local factors such as public health policies, population density, and mobility patterns play crucial roles in the spread of the virus (Anselin, 1995). These findings suggest that a one-size-fits-all approach may be insufficient in managing the pandemic, and that region-specific strategies are crucial.

In conclusion, while the ARIMA and ARIMAX models provided valuable tools for understanding and predicting COVID-19 trends, their limitations underscore the need for more complex models that can better capture the dynamic and non-linear nature of the pandemic (Gao et al., 2022). The multiple regression analysis highlighted the critical role of socioeconomic factors in determining COVID-19 case numbers, suggesting that public health interventions should be tailored to address these disparities. The spatial autocorrelation analysis further emphasized the importance of region-specific strategies in controlling the spread of the virus. Future research should focus on refining these models, incorporating more real-time data, and improving the granularity of spatial analyses to enhance their predictive power and applicability in public health decision-making. Additionally, the effect of vaccination on COVID-19 case numbers, as explored through various statistical techniques, highlights the critical role of timely and effective vaccination efforts in controlling the pandemic. However, the variability in outcomes across different regions suggests that a tailored, region-specific approach is essential for optimizing public health responses.

## Data Availability

All data produced in the present work are contained in the manuscript

## AUTHOR CONTRIBUTIONS

As the sole author, I was responsible for all aspects of this research, including the study design, data collection, data analysis, and manuscript preparation.

## ACKNOWLEDGMENTS

I would like to express my sincere gratitude to my supervisor, Dr. Wen Zhang, for his invaluable guidance, insightful feedback, and continuous support throughout the entire process of my research and thesis writing. His expertise, patience, and encouragement have been instrumental in the successful completion of this work.

## DATA AVAILABILITY STATEMENT

All datasets used in this study are listed in Table 1.

## FINANCIAL DISCLOSURE

None reported.

## CONFLICT OF INTEREST

The authors declare no potential conflict of interests.

## APPENDIX

### A EVALUATION PARAMETERS

Evaluating ARIMA models involves selecting the best model and measuring forecast accuracy using several key metrics. The most common evaluation criteria include Akaike Information Criterion (AIC), Bayesian Information Criterion (BIC), Root Mean Squared Error (RMSE), Mean Absolute Error (MAE), and Mean Squared Error (MSE). Each metric offers different insights into the model’s performance, making them suitable for various aspects of model evaluation (Hyndman & Athanasopoulos, 2018).

#### A.1 Akaike Information Criterion (AIC)

The Akaike Information Criterion (AIC) is widely used for model selection. It balances model fit and complexity by penalizing models with more parameters to avoid overfitting (Akaike, 1974). AIC is calculated as:

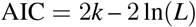

where *k* is the number of parameters in the model, and *L* is the likelihood of the model. A lower AIC value indicates a better model, as it reflects a good trade-off between model complexity and fit. However, AIC tends to favor slightly more complex models compared to BIC, as it imposes a lighter penalty on the number of parameters (Burnham & Anderson, 2004).

#### A.2 Bayesian Information Criterion (BIC)

The Bayesian Information Criterion (BIC) is another model selection criterion that penalizes model complexity more strongly than AIC (Schwarz, 1978). It is calculated as:

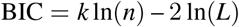

where *n* is the number of observations. Like AIC, a lower BIC value indicates a better model, but BIC tends to favor simpler models, particularly for larger datasets. BIC is more conservative than AIC, making it a better choice when the goal is to avoid overfitting and ensure generalizability to new data (Hyndman & Athanasopoulos, 2018).

#### A.3 Root Mean Squared Error (RMSE)

Root Mean Squared Error (RMSE) measures the average magnitude of the forecast errors by squaring the differences between the actual and predicted values before averaging them (Hyndman & Athanasopoulos, 2018). It is computed as:

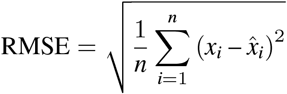

RMSE is sensitive to large errors due to the squaring of the residuals, making it particularly useful when larger deviations are more critical. However, this also means RMSE can be disproportionately affected by outliers (Chai & Draxler, 2014).

#### A.4 Mean Absolute Error (MAE)

Mean Absolute Error (MAE) measures the average magnitude of the errors without considering their direction. Unlike RMSE, MAE takes the absolute value of the errors, which makes it less sensitive to outliers (Willmott & Matsuura, 2005). MAE is calculated as:

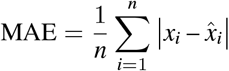

MAE provides a linear score, where all errors contribute equally to the metric, making it easier to interpret than RMSE. It is particularly useful when the focus is on the average error magnitude, regardless of the size of the deviations (Willmott & Matsuura, 2005).

#### A.5 Mean Squared Error (MSE)

Mean Squared Error (MSE) is another common metric that measures the average of the squared differences between the actual and predicted values. It is calculated as:

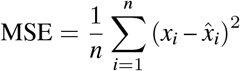

MSE, like RMSE, emphasizes larger errors due to the squaring of the residuals, but it does not take the square root, making it harder to interpret in the same units as the original data (Hyndman & Athanasopoulos, 2018). MSE is particularly useful when you want to penalize larger errors more heavily.

Each of these evaluation metrics offers unique characteristics that make them suitable for different scenarios. AIC and BIC focus on balancing model fit with complexity, with BIC being more conservative. RMSE and MSE are sensitive to larger errors, making them appropriate when outliers are important, while MAE provides a more robust measure against outliers by treating all errors equally. Depending on the objectives of the analysis, a combination of these metrics is often used to comprehensively assess and compare ARIMA models (Hyndman & Athanasopoulos, 2018).

### B MATHEMATICAL FORMULATIONS AND DETAILED ANALYSIS

#### B.1 Granger Causality Model Formulation

The Granger causality test was used to determine whether past values of the number of people vaccinated can help predict future values of new COVID-19 cases, suggesting a potential causal relationship. Formally, a time series *y*_*t*_ is said to be Granger-caused by another series *x*_*t*_ if past values of *x*_*t*_ provide statistically significant information about *y*_*t*_ in the presence of past values of *y*_*t*_ (Granger, 1969). The model can be represented as:

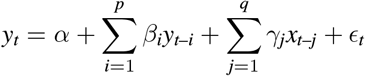

where:

- *y*_*t*_ represents the number of new COVID-19 cases,
- *x*_*t*_ represents the number of people vaccinated,
- *ϵ*_*t*_ is the error term,
- *α* is the intercept,
- *β*_*i*_ are the coefficients for the lagged values of *y*_*t*_,
- *γ*_*j*_ are the coefficients for the lagged values of *x*_*t*_.

The null hypothesis of the Granger causality test is that the coefficients *γ*_*j*_ are jointly zero, implying that *x*_*t*_ does not Granger-cause *y*_*t*_. In other words, if the null hypothesis is rejected, we conclude that past vaccination rates provide statistically significant predictive power for future COVID-19 cases.

#### B.2 Segmented Regression and Chow Test Formulation

The segmented regression model used to assess the impact of vaccination on COVID-19 case trends is expressed as:

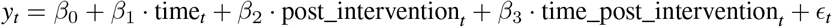

where:

- *y*_*t*_ is the number of new COVID-19 cases at time *t*,
- time_*t*_ is the time since the beginning of the study,
- post_intervention_*t*_ is a binary variable indicating whether the observation is post-intervention (e.g., after vaccination started),
- time_post_intervention_*t*_ is the time since the intervention began,
- *ϵ*_*t*_ is the error term.

The coefficients of interest are:

- *β*_2_: Represents the immediate change in level after the intervention,
- *β*_3_: Represents the change in trend following the intervention.

To further validate the segmented regression, a Chow test was performed to detect any structural breaks at the point of intervention. The test compares the sum of squared residuals (SSR) from three different models:

1. The full model (including all data),
2. The pre-intervention model (data before vaccination),
3. The post-intervention model (data after vaccination).

The test statistic is given by:

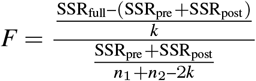

where:

- SSR_full_ is the sum of squared residuals from the full model,
- SSR_pre_ and SSR_post_ are the sums of squared residuals from the pre-intervention and post-intervention models, respectively,
- *k* is the number of parameters in the model,
- *n*_1_ and *n*_2_ are the number of observations before and after the intervention, respectively.

The null hypothesis of the Chow test states that there is no structural break at the intervention point, implying that the coefficients remain consistent before and after the intervention. Rejecting the null hypothesis indicates a significant structural break, suggesting that the intervention (e.g., vaccination) caused a change in the trend of new COVID-19 cases.

#### B.2 Regression Discontinuity Design (RDD) Model Formulation

The RDD model used to estimate the causal effect of vaccine introduction on new COVID-19 cases is expressed as:

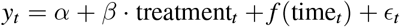

where:

- *y*_*t*_: Represents the number of new COVID-19 cases at time *t*,
- treatment_*t*_: An indicator variable equal to 1 if the observation occurred after the cutoff (e.g., after the start of mass vaccination), and 0 otherwise,
- *f* (time_*t*_): A smooth function of time, allowing for flexibility in the time trend on either side of the cutoff,
- *α*: The intercept term,
- *β*: The coefficient that represents the treatment effect of vaccination at the cutoff point,
- *ϵ*_*t*_: The error term.

The RDD approach relies on the assumption that observations close to the cutoff point are comparable, except for the treatment effect induced by the introduction of vaccines. The non-parametric approach used in this study allows for a flexible functional form for *f* (time_*t*_), avoiding restrictive assumptions about the relationship between time and new COVID-19 cases on either side of the cutoff (Lee & Lemieux, 2010).

#### B.4 Regression Model and Correlation Analysis Formulation

The linear regression model used to investigate the relationship between COVID-19 infection rates and economic development is formulated as follows:

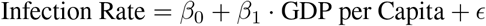

where:

- *β*_0_ is the intercept,
- *β*_1_ is the coefficient for GDP per capita,
- *ϵ* is the error term.

In addition to the regression analysis, Pearson, Spearman, and Maximal Information Coefficient (MIC) were calculated to further measure the strength and direction of the association between GDP per capita and COVID-19 infection rates. The Pearson correlation coefficient is calculated as:

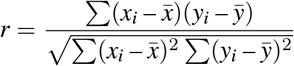

The Spearman rank correlation is defined as the Pearson correlation between the ranked variables. Spearman’s coefficient assesses the strength of a monotonic relationship between two variables.

Additionally, the Maximal Information Coefficient (MIC) was computed to capture any potential nonlinear relationships between GDP per capita and COVID-19 infection rates. MIC is based on mutual information and measures the strength of the association between two variables without assuming a linear relationship. It is designed to detect both linear and nonlinear dependencies, and its value ranges from 0 (no association) to 1 (perfect association).

#### B.5 Expanded Multiple Regression Model Formulation

The expanded multiple regression model used to investigate the determinants of COVID-19 infection rates is specified as follows:

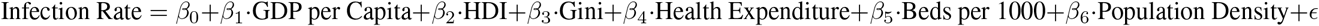

where:

- *β*_0_: The intercept,
- *β*_1_ · GDP per Capita: Coefficient for GDP per capita,
- *β*_2_ · HDI: Coefficient for Human Development Index (HDI),
- *β*_3_ · Gini: Coefficient for Gini coefficient,
- *β*_4_ · Health Expenditure: Coefficient for per capita health expenditure,
- *β*_5_ · Beds per 1000: Coefficient for hospital beds per 1,000 people,
- *β*_6_ · Population Density: Coefficient for population density,
- *ϵ*: Error term.

Interaction terms were included to investigate the potential synergistic effects between these variables. For instance, interaction between health expenditure and GDP per capita was examined to understand how healthcare investment may influence the relationship between economic development and infection rates. Additionally, interactions between population density and other socioeconomic factors were analyzed to assess the impact of urbanization on infection spread (Montgomery et al., 2012).

#### B.6 Principal Component Regression (PCR) and Partial Least Squares (PLS) Regression Formulation

Principal Component Regression (PCR) involves performing Principal Component Analysis (PCA) on the predictor variables and then using the principal components as predictors in the regression model. The PCR model is formulated as follows:

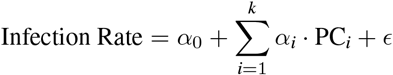

where:

- *α*_0_: The intercept term,
- PC_*i*_: The *i*-th principal component extracted from the predictor variables,
- *α*_*i*_: The coefficient corresponding to the *i*-th principal component,
- *k*: The number of principal components included in the model,
- *ϵ*: The error term.

Principal components are uncorrelated, and the first few components capture the maximum variance in the predictor variables. Cross-validation was used to determine the optimal number of components to include in the model, balancing model complexity and prediction accuracy (Jolliffe, 2002).

Partial Least Squares (PLS) regression is similar to PCR but extends the approach by considering the covariance between the predictors and the dependent variable when determining the components. The PLS model is written similarly to PCR, but typically requires fewer components because it selects components that are more directly related to the dependent variable (Wold et al., 2001). The PLS model can be expressed as:

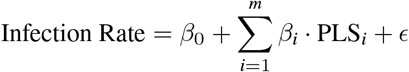

where:

- *β*_0_: The intercept term,
- PLS_*i*_: The *i*-th PLS component,
- *β*_*i*_: The coefficient corresponding to the *i*-th PLS component,
- *m*: The number of PLS components included in the model.

Similar to PCR, cross-validation was performed to determine the optimal number of components for PLS. The models were evaluated based on the Mean Squared Error of Prediction (MSEP), and component loadings were analyzed to interpret the contribution of the original variables to the extracted components (Jolliffe & Cadima, 2016).

#### B.7 Mathematical Formulation of Moran’s I

Moran’s I is a measure of global spatial autocorrelation that quantifies the degree of spatial clustering in a variable across geographic space. Mathematically, Moran’s I is expressed as:

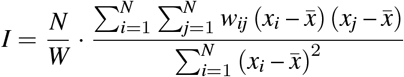

where:

- *N* is the total number of spatial units (e.g., states or regions),
- *x*_*i*_ and *x*_*j*_ are the values of the variable of interest (e.g., COVID-19 infection rates) at locations *i* and *j*,
- 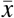 is the mean value of the variable,
- *w*_*ij*_ is the spatial weight between locations *i* and *j*, indicating the strength of the spatial relationship (e.g., based on shared boundaries),
- *W* is the sum of all spatial weights, i.e., 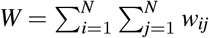 (Cliff & Ord, 1981).

Moran’s I ranges from -1 to 1, where:

- *I* > 0: Indicates positive spatial autocorrelation, meaning similar values are spatially clustered.
- *I* < 0: Indicates negative spatial autocorrelation, meaning dissimilar values are adjacent.
- *I* = 0: Suggests a random spatial distribution of values.

For this analysis, the spatial weights matrix was generated based on shared boundaries between geographic regions. Moran’s I was computed to assess the overall spatial autocorrelation of COVID-19 infection rates, using boundary-based spatial relationships to understand the clustering behavior of infection rates (Anselin, 1995).

#### B.8 Mathematical Formulation of the Getis-Ord Gi* Statistic

The Getis-Ord Gi* (G-star) statistic is a local spatial statistic used to identify hotspots (areas of high-value clustering) and coldspots (areas of low-value clustering) within a geographic region. The Gi^*\**^ statistic for a location *i* is calculated as:

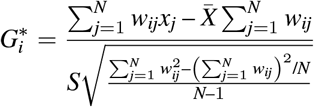

where:

- *x*_*j*_: Represents the value of the variable of interest (e.g., COVID-19 infection rates) at location *j*,
- 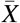: The mean value of the variable across all locations,
- *S*: The standard deviation of the variable,
- *w*_*ij*_: The spatial weight between locations *i* and *j*, indicating the strength of their spatial relationship,
- *N*: The total number of spatial units (e.g., regions or states) (Ord & Getis, 1995).

A significantly positive 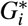 value indicates the presence of a hotspot (i.e., clustering of high values), while a significantly negative 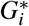 value indicates a coldspot (i.e., clustering of low values). The significance of the 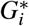 values is determined by comparing the observed statistic to a reference distribution under the null hypothesis of spatial randomness (Getis & Ord, 1992). For this analysis, the same spatial weights matrix was used to compute the Getis-Ord Gi* statistic, identifying geographic regions with significant clustering of COVID-19 infection rates. These regions were classified as hotspots or coldspots depending on the sign and significance of the 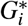 statistic.

### C DETECTED OUTLIERS IN GLOBAL AND USA COVID-19 CASES

**FIGURE C1.**
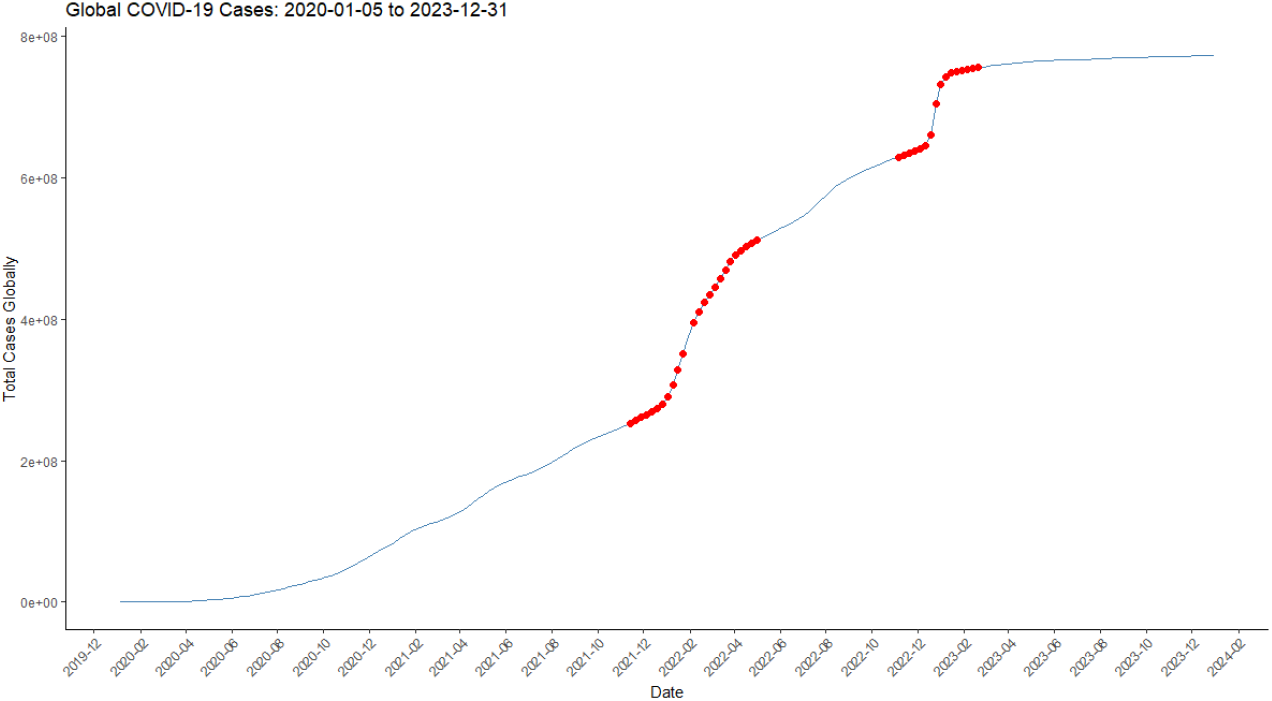
Global COVID-19 Cases outliers detected: 2020-01-05 to 2023-12-31

**FIGURE C2.**
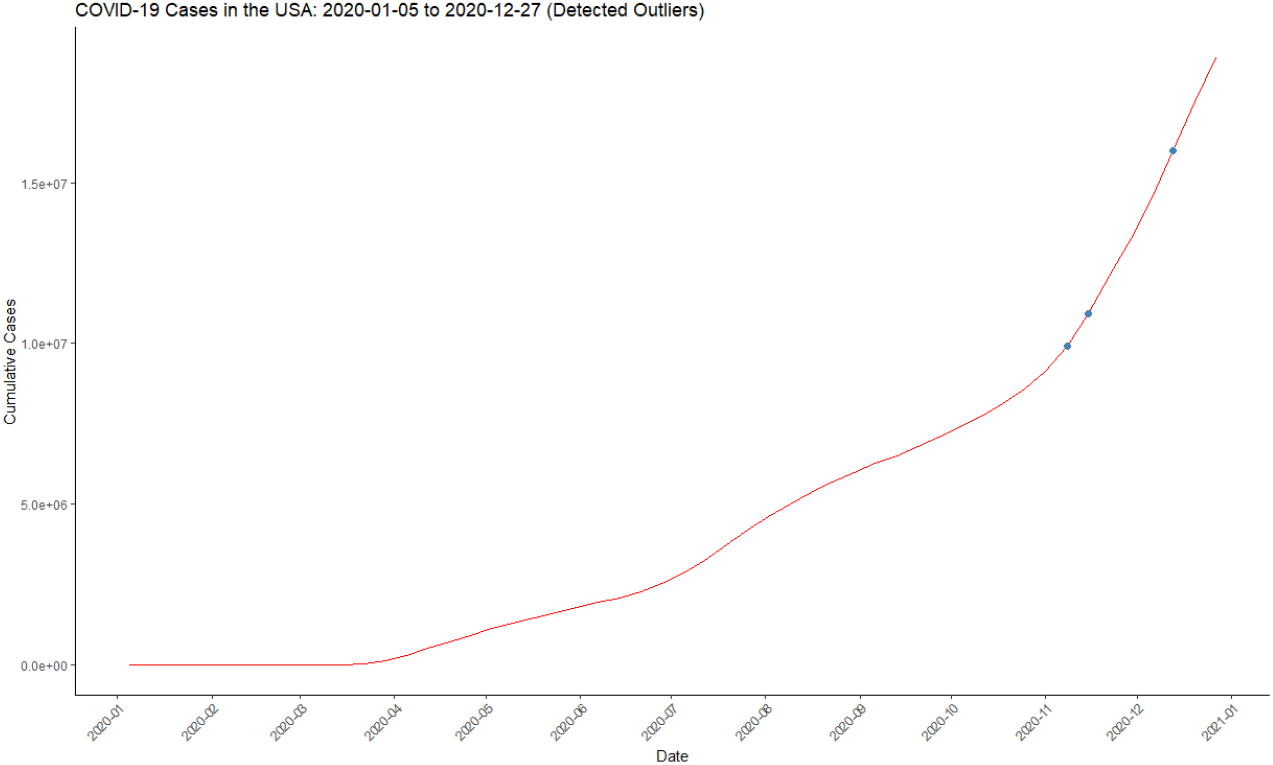
COVID-19 Cases in the USA: 2020-01-05 to 2020-12-27 (Detected Outliers)

**TABLE C1.** Outliers detected points of USA and Global.

| (a) Outliers detected points of USA |  | (b) Outliers detected points of Global |  |
| --- | --- | --- | --- |
| Date | Cumulative cases | Date | Cumulative cases |
| 2021-11-07 | 46149896 | 2021-11-14 | 253323118 |
| 2021-11-14 | 46707076 | 2021-11-21 | 257186504 |
| 2021-11-21 | 47383952 | 2021-11-28 | 261233860 |
| 2021-11-28 | 47976346 | 2021-12-05 | 265507253 |
| 2021-12-05 | 48727363 | 2021-12-12 | 269821260 |
| 2021-12-12 | 49562332 | 2021-12-19 | 274524087 |
| 2021-12-19 | 50461441 | 2021-12-26 | 280614414 |
| 2021-12-26 | 51878860 | 2022-01-02 | 291083242 |
| 2022-01-02 | 54590898 | 2022-01-09 | 307680730 |
| 2022-01-09 | 59273795 | 2022-01-16 | 328342435 |
| 2022-01-23 | 69967616 | 2022-01-23 | 351884049 |
| 2022-01-30 | 73756878 | 2022-02-06 | 395236775 |
| 2022-02-06 | 75945588 | 2022-02-13 | 411148680 |
| 2022-02-13 | 77167700 | 2022-02-20 | 424020354 |
| 2022-02-20 | 77875856 | 2022-02-27 | 435070227 |
| 2022-02-27 | 78334128 | 2022-03-06 | 445970477 |
| 2022-03-06 | 78666637 | 2022-03-13 | 457775947 |
| 2022-03-13 | 78900969 | 2022-03-20 | 470154333 |
| 2022-03-20 | 79109709 | 2022-03-27 | 481152614 |
|  |  | 2022-04-03 | 490521795 |
|  |  | 2022-04-10 | 497721977 |
|  |  | 2022-04-17 | 503419788 |
|  |  | 2022-04-24 | 508123035 |
|  |  | 2022-05-01 | 512121948 |
|  |  | 2022-11-06 | 629416831 |
|  |  | 2022-11-13 | 631906011 |
|  |  | 2022-11-20 | 634588061 |
|  |  | 2022-11-27 | 637565688 |
|  |  | 2022-12-04 | 640802697 |
|  |  | 2022-12-11 | 645417862 |
|  |  | 2022-12-18 | 660395376 |
|  |  | 2022-12-25 | 704631603 |
|  |  | 2023-01-01 | 732364303 |
|  |  | 2023-01-08 | 743344318 |
|  |  | 2023-01-15 | 748231266 |
|  |  | 2023-01-22 | 750746154 |
|  |  | 2023-01-29 | 752305655 |
|  |  | 2023-02-05 | 753691312 |
|  |  | 2023-02-12 | 754938671 |
|  |  | 2023-02-19 | 756084464 |

### D ARIMA MODEL ANALYSIS ACROSS DIFFERENT CONTINENTS

**FIGURE D3.**
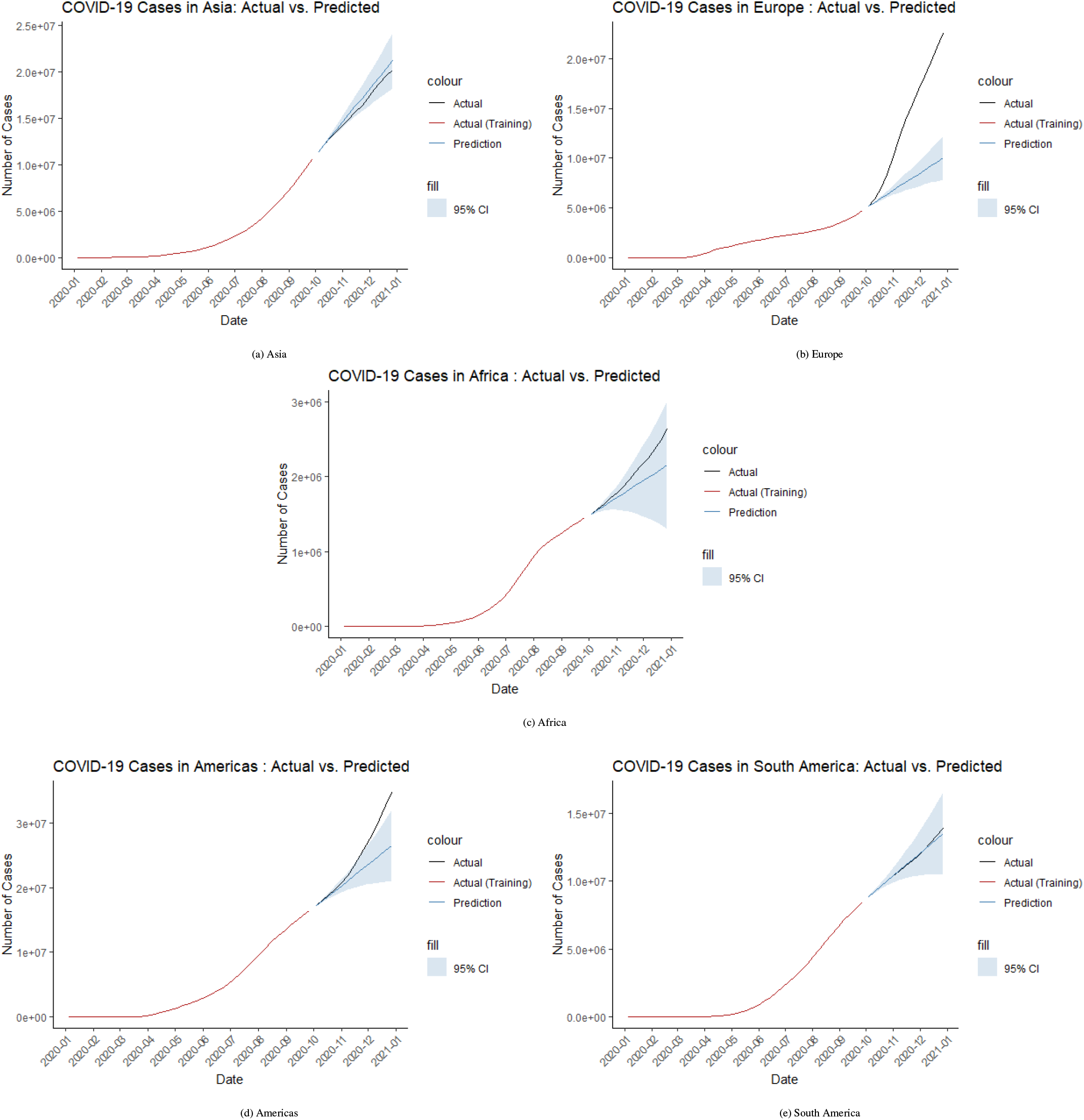
ARIMA Model Analysis across different Continents

### E RMSE COMPARISON AND FORECAST ANALYSIS FOR ARIMA MODELS (EUROPE)

**TABLE E2.** RMSE Comparison between auto.arima and Cross-Validation for ARIMA Models (Europe).

| Model | ARIMA Parameters |  |  | RMSE |
| --- | --- | --- | --- | --- |
|  | p | d | q | RMSE |
| auto.arima | 0 | 2 | 2 | 14479.46 |
| Cross-Validation | 1 | 2 | 0 | 9981.767 |

**FIGURE E4.**
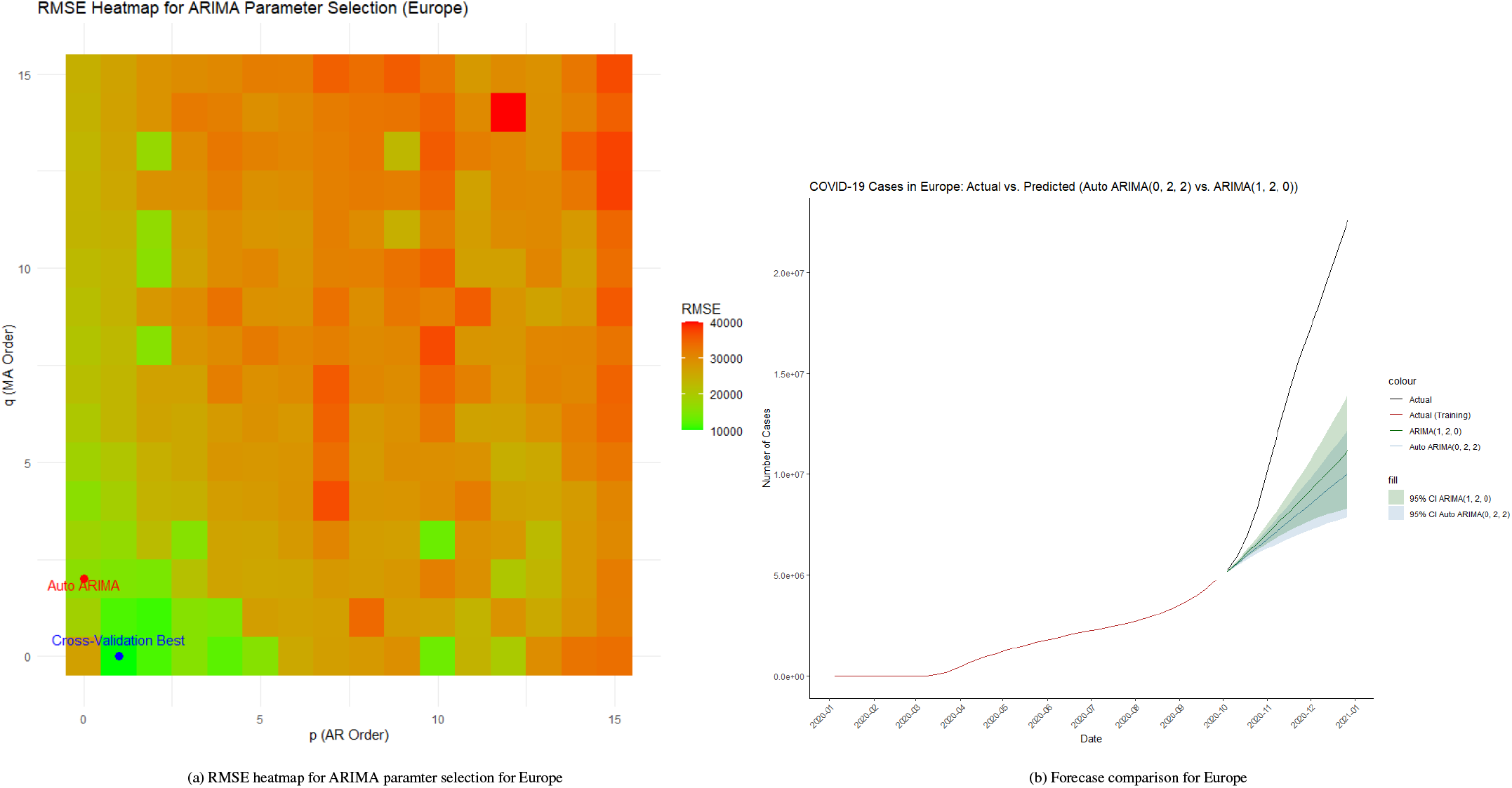
Heatmap of RMSE and forecast comparison of Europe

### F DETAILED RESULTS OF SEGMENTED REGRESSION AND REGRESSION DISCONTINUITY

**TABLE F3.** Segmented Regression Output.

| Coefficients | Estimate | Std. Error | t value | Pr(>\|t\|) |
| --- | --- | --- | --- | --- |
| (Intercept) | -111033 | 213511 | -0.520 | 0.60372 |
| time | 18987 | 8173 | 2.323 | 0.02136* |
| post_intervention | 260639 | 255961 | 1.018 | 0.31000 |
| time_post_intervention | -24115 | 8365 | -2.883 | 0.00445** |
| Residuals: |  |  |  |  |
| Min: -803922, 1Q: -274147, Median: -105598, 3Q: 81517, Max: 4920221 |  |  |  |  |
| Residual standard error: 735900 on 169 degrees of freedom |  |  |  |  |
| Multiple R-squared: 0.1192 |  |  |  |  |
| Adjusted R-squared: 0.1036 |  |  |  |  |
| F-statistic: 7.623 on 3 and 169 DF, p-value: 8.242e-05 |  |  |  |  |
| Signif. codes: 0 '***' 0.001 '**' 0.01 '*' 0.05 '.' 0.1 ' ' 1 |  |  |  |  |

**TABLE F4.** Regression Discontinuity.

|  | Left of Cutoff |  | Right of Cutoff |
| --- | --- | --- | --- |
| Number of Obs. | 46 |  | 127 |
| BW type |  | mserd |  |
| Kernel |  | Triangular |  |
| VCE method |  | NN |  |
| Eff. Number of Obs. | 5 |  | 6 |
| Order est. (p) | 1 |  | 1 |
| Order bias (q) | 2 |  | 2 |
| BW est. (h) | 5.520 |  | 5.520 |
| BW bias (b) | 9.645 |  | 9.645 |
| rho (h/b) | 0.572 |  | 0.572 |
| Unique Obs. | 46 |  | 127 |

| Method | Coef. | Std. Err. | z | P>\|z\| |
| --- | --- | --- | --- | --- |
| [95% C.I.] |  |  |  |  |
| Conventional | 76662.15 | 162147.38 | 0.473 | 0.636 |
| [-241140.87 , 394465.18] |  |  |  |  |
| Robust | - | - | 0.387 | 0.699 |
| [-315644.73 , 470854.24] |  |  |  |  |

### G DETAILED RESULTS OF ARIMAX MODEL

**TABLE G5.**
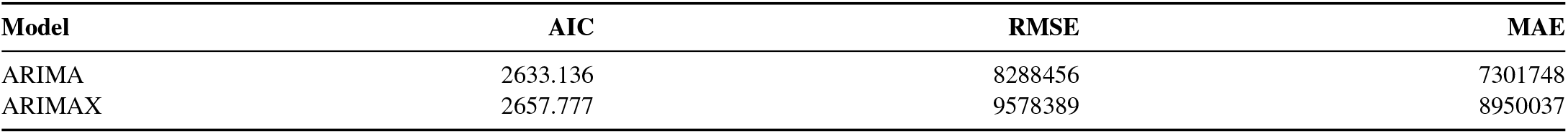
Comparison of ARIMA and ARIMAX Models (Period 2).

### H TOP 10 COUNTRIES BY COVID-19 INFECTION RATE ON 2023-12-31

**FIGURE G5.**
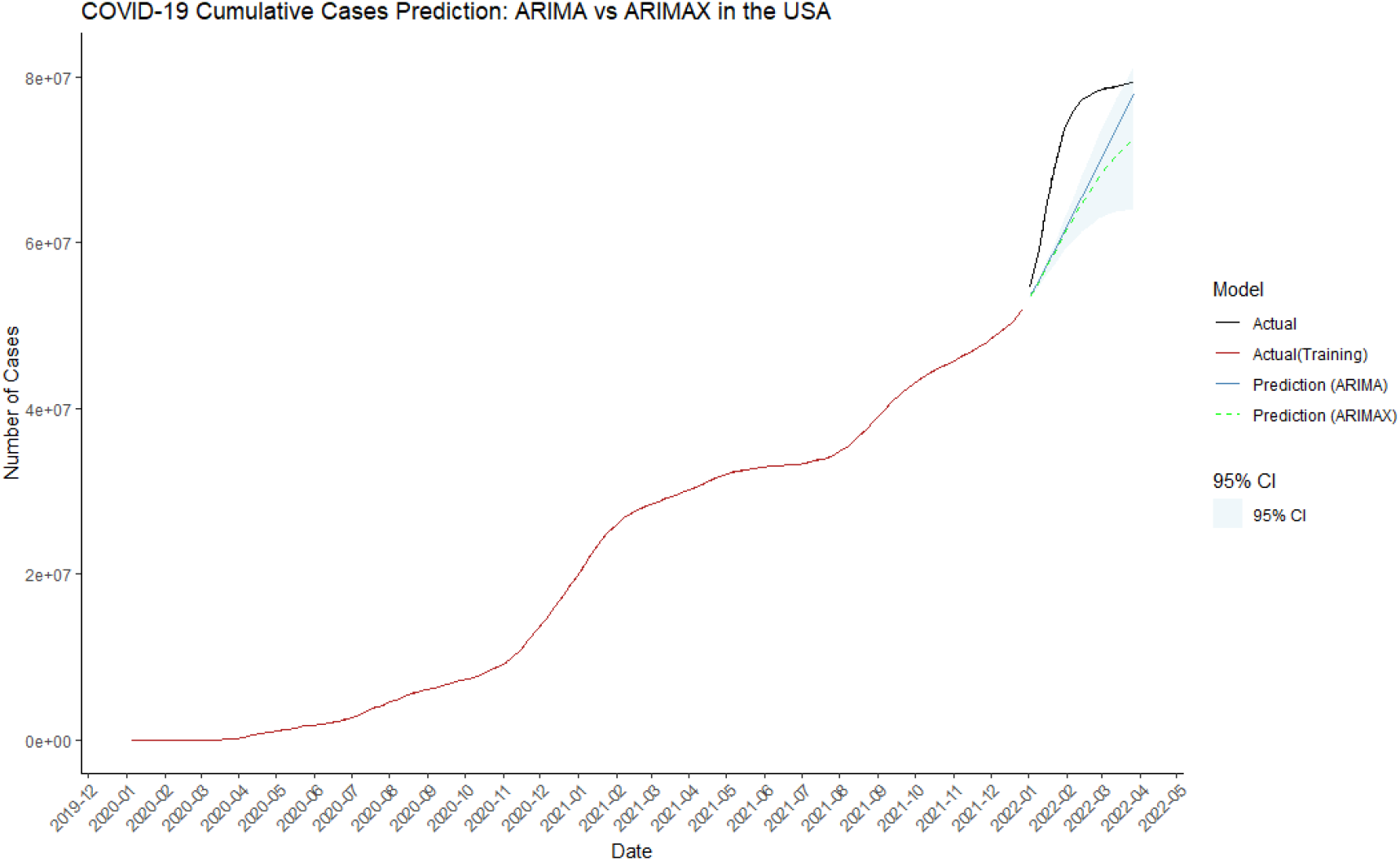
ARIMAX forecast for period 2

**FIGURE G6.**
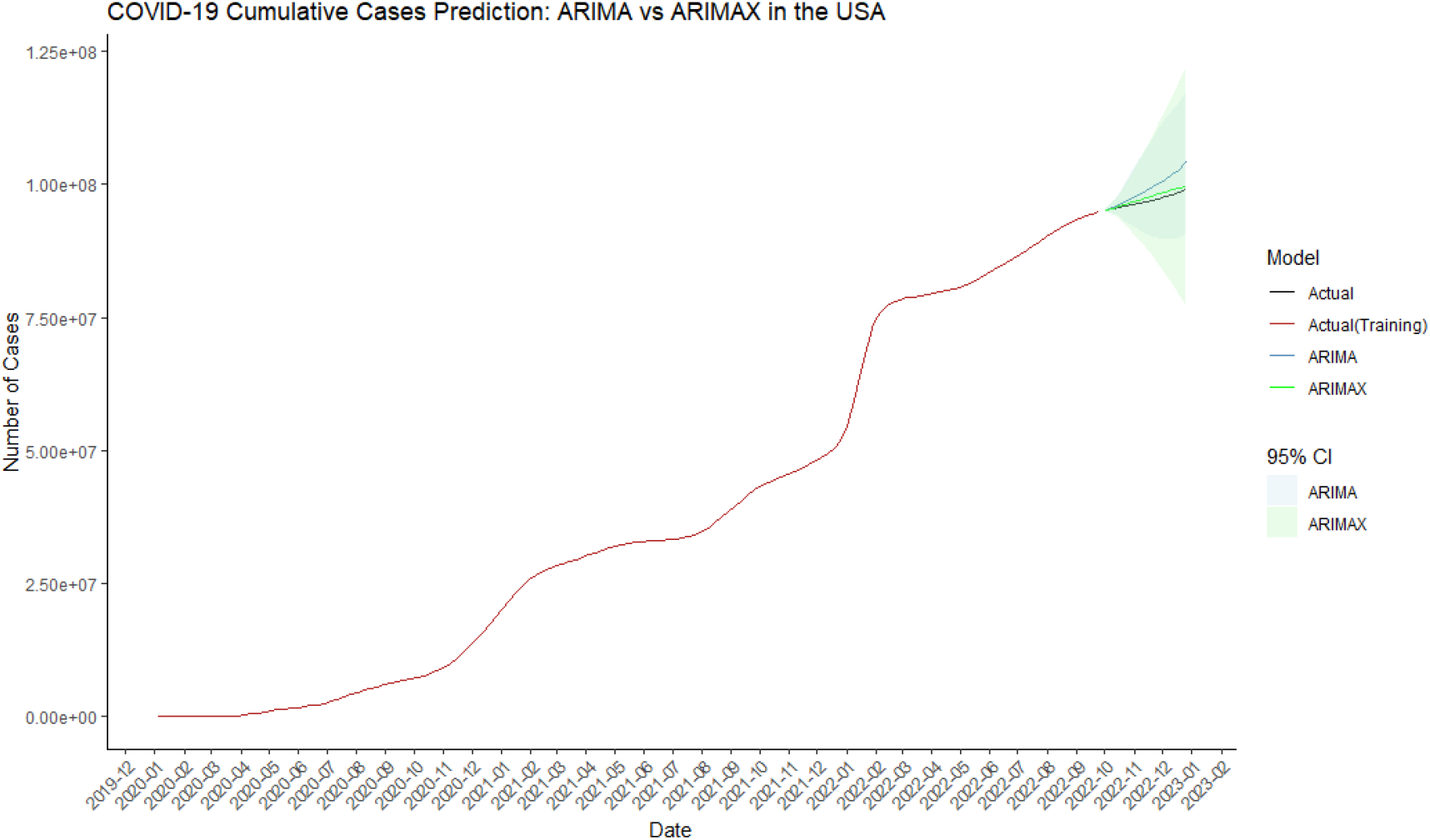
ARIMAX forecast for period 3

**TABLE G6.**
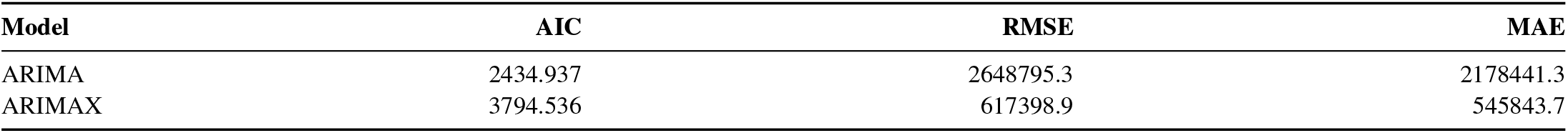
Comparison of ARIMA and ARIMAX Models (Period 3).

**FIGURE H7.**
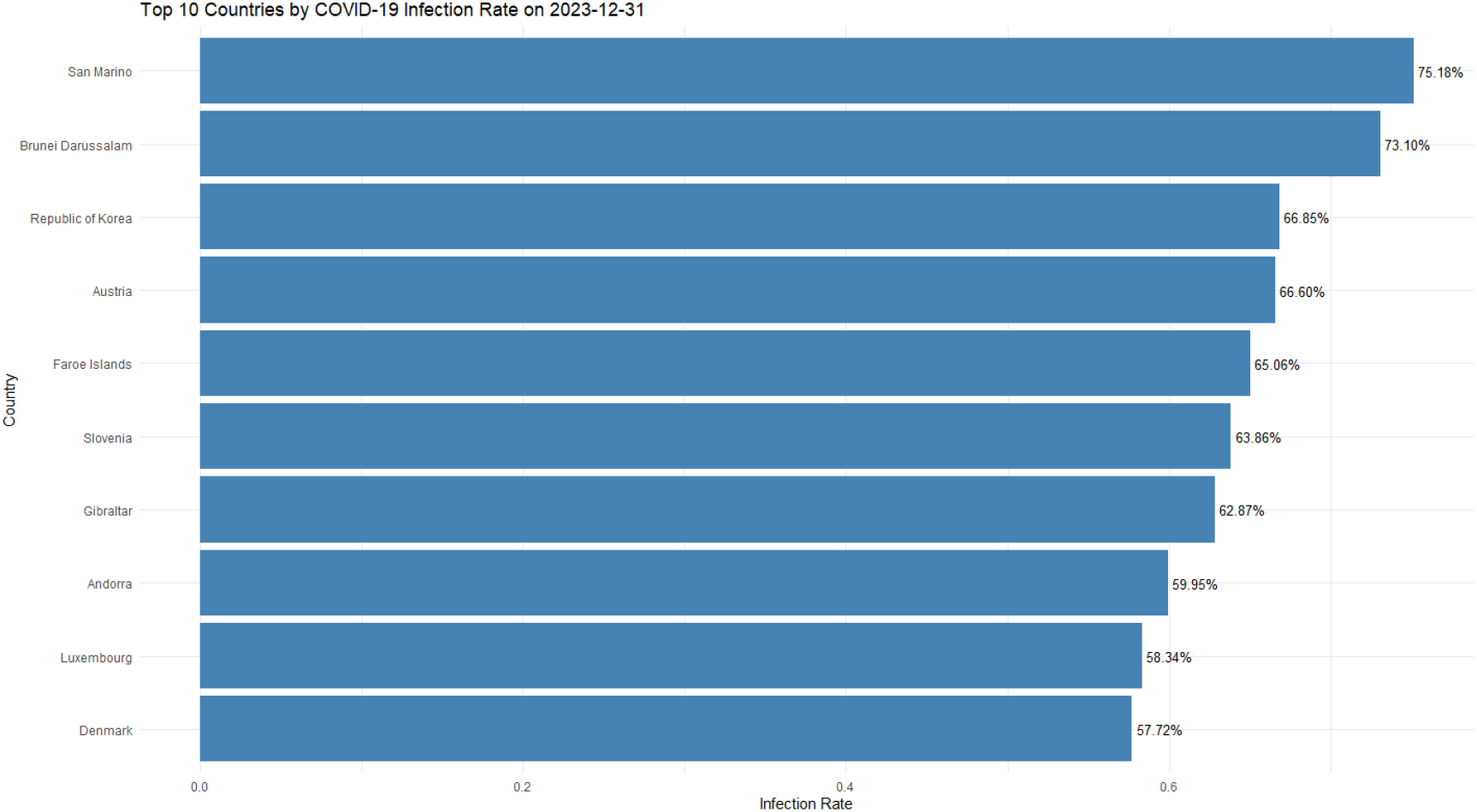
Top 10 Countries by COVID-19 Infection Rate on 2023-12-31

### I MULTIPLE REGRESSION RESULTS

**TABLE I7.** Multiple Regression Results: Infection Rate vs. Socioeconomic and Health Factors.

| Coefficients | Estimate | Std. Error | t value | Pr(>\|t\|) |
| --- | --- | --- | --- | --- |
| (Intercept) | -1.983e+00 | 1.466e+00 | -1.353 | 0.183801 |
| GDP per capita | -2.556e-04 | 8.909e-05 | -2.869 | 0.006544 ** |
| HDI | 3.305e+00 | 2.095e+00 | 1.578 | 0.122523 |
| Gini | 5.595e-02 | 4.314e-02 | 1.297 | 0.202078 |
| Health_Expenditure | 3.226e-03 | 9.324e-04 | 3.460 | 0.001297 ** |
| Beds_per_1000 | -1.624e-01 | 1.589e-01 | -1.022 | 0.312785 |
| Population_Density | -1.753e-03 | 2.132e-03 | -0.822 | 0.416031 |
| GDP per capita:HDI | 1.937e-04 | 6.730e-05 | 2.879 | 0.006381 ** |
| GDP per capita:Gini | 2.490e-06 | 1.104e-06 | 2.255 | 0.029676 * |
| GDP per capita:Health_Expenditure | 9.444e-10 | 5.735e-10 | 1.647 | 0.107478 |
| GDP per capita:Beds_per_1000 | -2.560e-06 | 2.457e-06 | -1.042 | 0.303709 |
| GDP per capita:Population_Density | -5.759e-09 | 2.454e-08 | -0.235 | 0.815648 |
| HDI:Gini | -8.932e-02 | 6.185e-02 | -1.444 | 0.156496 |
| HDI:Health_Expenditure | -2.918e-03 | 7.967e-04 | -3.662 | 0.000724 *** |
| HDI:Beds_per_1000 | 3.109e-01 | 1.936e-01 | 1.606 | 0.116167 |
| HDI:Population_Density | 3.461e-03 | 2.981e-03 | 1.161 | 0.252460 |
| Gini:Health_Expenditure | -1.901e-05 | 8.145e-06 | -2.333 | 0.024737 * |
| Gini:Beds_per_1000 | -1.251e-03 | 1.789e-03 | -0.699 | 0.488522 |
| Gini:Population_Density | -1.178e-05 | 3.779e-05 | -0.312 | 0.756886 |
| Health_Expenditure:Beds_per_1000 | 1.944e-05 | 1.622e-05 | 1.198 | 0.237825 |
| Health_Expenditure:Population_Density | 5.495e-08 | 2.320e-07 | 0.237 | 0.813981 |
| Beds_per_1000:Population_Density | -2.616e-04 | 1.166e-04 | -2.244 | 0.030425 * |

### J COEFFICIENT PLOT FOR MULTIPLE REGRESSION ANALYSIS

**FIGURE J8.**
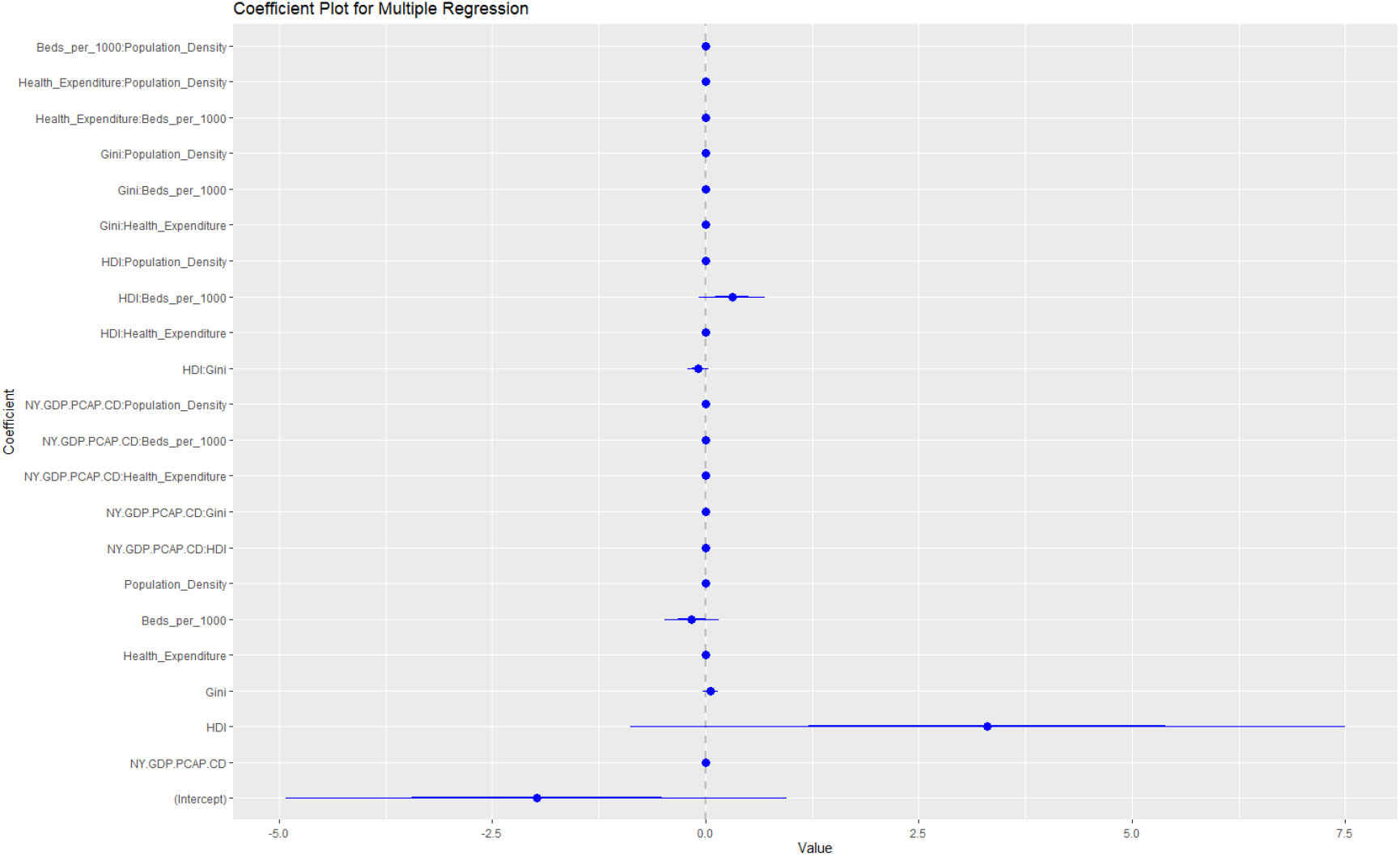
Coefficient Plot for the Multiple Regression Analysis

### K PCR RESULTS AND CROSS-VALIDATION RESULTS: RMSEP VALUES

**TABLE K8.**
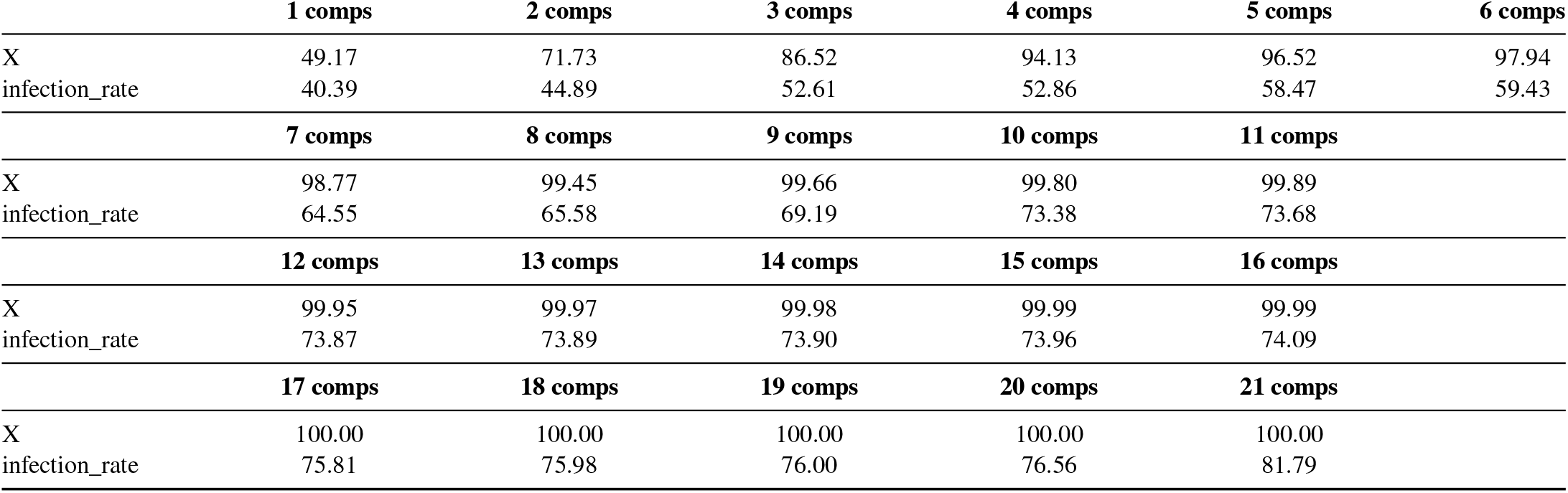
PCR Results: Variance Explained by Number of Components.

|  | 1 comps | 2 comps | 3 comps | 4 comps | 5 comps | 6 comps |
| --- | --- | --- | --- | --- | --- | --- |
| X | 49.17 | 71.73 | 86.52 | 94.13 | 96.52 | 97.94 |
| infection_rate | 40.39 | 44.89 | 52.61 | 52.86 | 58.47 | 59.43 |
|  | 7 comps | 8 comps | 9 comps | 10 comps | 11 comps |  |
| X | 98.77 | 99.45 | 99.66 | 99.80 | 99.89 |  |
| infection_rate | 64.55 | 65.58 | 69.19 | 73.38 | 73.68 |  |
|  | 12 comps | 13 comps | 14 comps | 15 comps | 16 comps |  |
| X | 99.95 | 99.97 | 99.98 | 99.99 | 99.99 |  |
| infection_rate | 73.87 | 73.89 | 73.90 | 73.96 | 74.09 |  |
|  | 17 comps | 18 comps | 19 comps | 20 comps | 21 comps |  |
| X | 100.00 | 100.00 | 100.00 | 100.00 | 100.00 |  |
| infection_rate | 75.81 | 75.98 | 76.00 | 76.56 | 81.79 |  |

### L PLS LOADINGS AND FINAL REGRESSION COEFFICIENTS

**TABLE K9.** Cross-Validation Results.

|  | (Intercept) | 1 comps | 2 comps | 3 comps | 4 comps | 5 comps |
| --- | --- | --- | --- | --- | --- | --- |
| CV | 0.1868 | 0.1430 | 0.1331 | 0.1416 | 0.1253 | 0.1158 |
| adjCV | 0.1868 | 0.1423 | 0.1325 | 0.1402 | 0.1242 | 0.1159 |
|  | 6 comps | 7 comps | 8 comps | 9 comps | 10 comps | 11 comps |
| CV | 0.1309 | 0.1313 | 0.1266 | 0.1243 | 0.1377 | 0.1601 |
| adjCV | 0.1293 | 0.1290 | 0.1246 | 0.1225 | 0.1348 | 0.1558 |
|  | 12 comps | 13 comps | 14 comps | 15 comps | 16 comps | 17 comps |
| CV | 0.1791 | 0.1791 | 0.1905 | 0.2013 | 0.2269 | 0.2478 |
| adjCV | 0.1733 | 0.1734 | 0.1841 | 0.1943 | 0.2184 | 0.2379 |
|  | 18 comps | 19 comps | 20 comps | 21 comps |  |  |
| CV | 0.2568 | 0.2265 | 0.1950 | 0.2016 |  |  |
| adjCV | 0.2464 | 0.2179 | 0.1877 | 0.1941 |  |  |

**TABLE L10.** PLS Loadings.

|  | Comp 1 | Comp 2 | Comp 3 | Comp 4 | Comp 5 |
| --- | --- | --- | --- | --- | --- |
| NY.GDP.PCAP.CD | 0.297 |  | -0.200 | 0.171 |  |
| HDI | 0.263 | 0.242 | 0.366 |  | -0.218 |
| Gini | -0.172 | -0.106 | -0.174 |  | -0.840 |
| Health_Expenditure | 0.293 |  | -0.255 | 0.130 |  |
| Beds_per_1000 | 0.160 | 0.364 | 0.320 | -0.375 |  |
| Population_Density |  | -0.386 | 0.518 | -0.265 |  |
| NY.GDP.PCAP.CD:HDI | 0.297 |  | -0.206 | 0.170 |  |
| NY.GDP.PCAP.CD:Gini | 0.289 | -0.100 | -0.240 | 0.172 | -0.115 |
| NY.GDP.PCAP.CD:Health_Expenditure | 0.274 | -0.128 | -0.384 |  |  |
| NY.GDP.PCAP.CD:Beds_per_1000 | 0.306 |  |  |  | 0.125 |
| NY.GDP.PCAP.CD:Population_Density | 0.186 | -0.336 | 0.407 | -0.117 |  |
| HDI:Gini |  |  | 0.365 |  | -1.086 |
| HDI:Health_Expenditure | 0.293 |  | -0.259 | 0.129 |  |
| HDI:Beds_per_1000 | 0.192 | 0.341 | 0.316 | -0.321 |  |
| HDI:Population_Density |  | -0.375 | 0.536 | -0.241 |  |
| Gini:Health_Expenditure | 0.282 | -0.104 | -0.298 | 0.115 | -0.175 |
| Gini:Beds_per_1000 | 0.128 | 0.365 | 0.287 | -0.388 | -0.345 |
| Gini:Population_Density |  | -0.399 | 0.494 | -0.270 | -0.105 |
| Health_Expenditure:Beds_per_1000 | 0.298 |  | -0.122 |  |  |
| Health_Expenditure:Population_Density | 0.188 | -0.330 | 0.414 | -0.126 |  |
| Beds_per_1000:Population_Density | 0.125 | -0.280 | 0.569 | -0.321 |  |

### M PLS MODEL COMPONENT ANALYSIS

### N STATE-LEVEL COVID-19 CASES AND HOTSPOT/COLDSPOT ANALYSIS

**TABLE L11.** Final Coefficients (infection_rate)

| Variable | Coefficient |
| --- | --- |
| NY.GDP.PCAP.CD | 0.013031859 |
| HDI | 0.079431890 |
| Gini | -0.044192005 |
| Health_Expenditure | -0.004507819 |
| Beds_per_1000 | 0.004341445 |
| Population_Density | -0.019700953 |
| NY.GDP.PCAP.CD:HDI | 0.009171864 |
| NY.GDP.PCAP.CD:Gini | 0.014297801 |
| NY.GDP.PCAP.CD:Health_Expenditure | -0.060740881 |
| NY.GDP.PCAP.CD:Beds_per_1000 | 0.027603250 |
| NY.GDP.PCAP.CD:Population_Density | 0.043032791 |
| HDI:Gini | 0.012604470 |
| HDI:Health_Expenditure | -0.009862007 |
| HDI:Beds_per_1000 | 0.023099882 |
| HDI:Population_Density | -0.009237630 |
| Gini:Health_Expenditure | -0.006661809 |
| Gini:Beds_per_1000 | -0.016399951 |
| Gini:Population_Density | -0.026722755 |
| Health_Expenditure:Beds_per_1000 | 0.022314704 |
| Health_Expenditure:Population_Density | 0.040426417 |
| Beds_per_1000:Population_Density | -0.036921576 |

**FIGURE M9.**
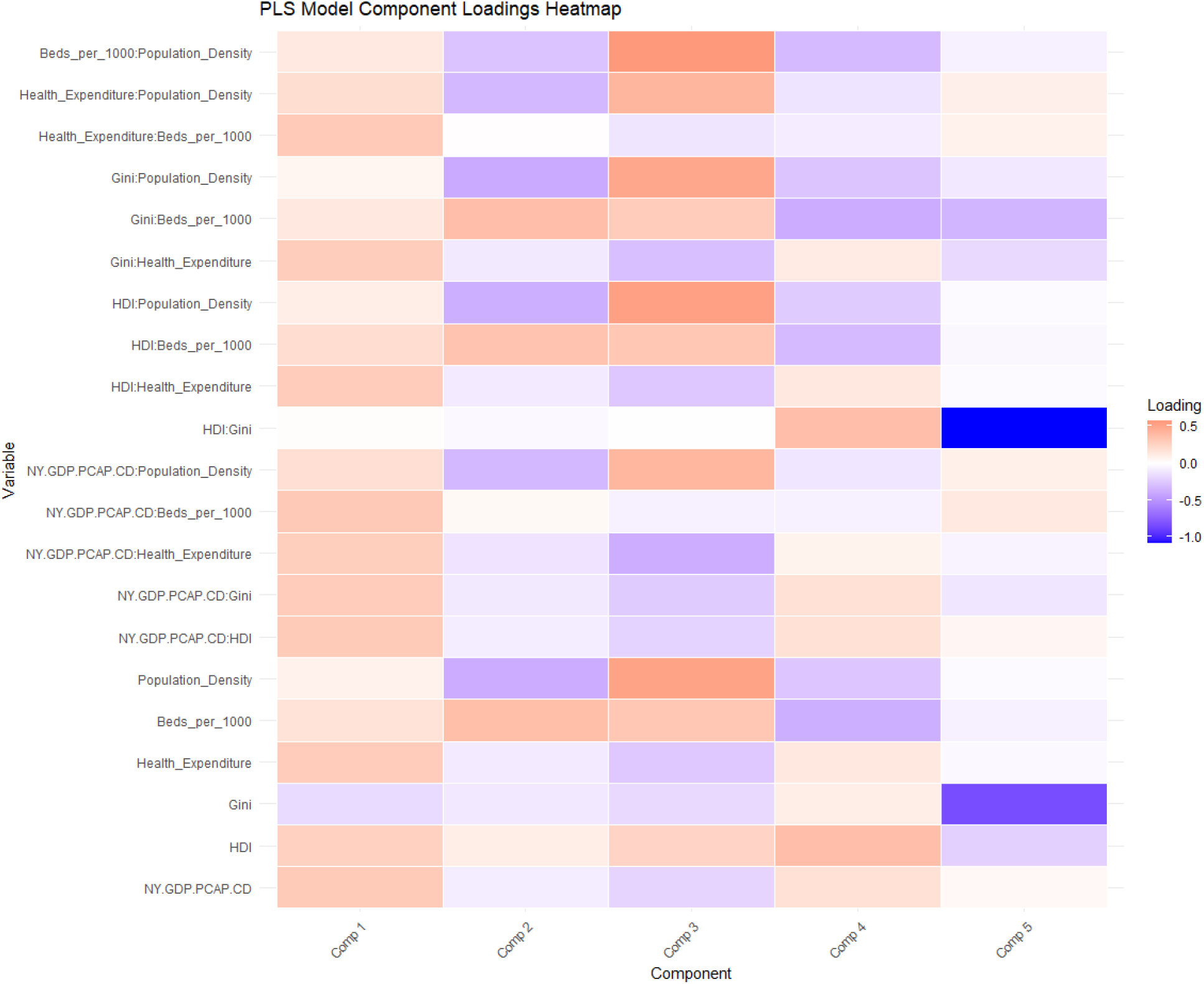
PLS Model Component Loadings Heatmap

**TABLE N12.** Top 10 and Bottom 10 States by COVID-19 Cases per 100,000 People.

| Rank | Top 10 States | Cases per 100,000 | Rank | Bottom 10 States | Cases per 100,000 |
| --- | --- | --- | --- | --- | --- |
| 1 | Alaska | 40,576.16 | 1 | New York | 18,251.51 |
| 2 | Rhode Island | 40,281.14 | 2 | Maryland | 22,319.23 |
| 3 | Kentucky | 38,512.10 | 3 | Oregon | 23,051.58 |
| 4 | North Dakota | 37,132.71 | 4 | Maine | 23,140.85 |
| 5 | West Virginia | 36,753.10 | 5 | Vermont | 23,822.64 |
| 6 | Tennessee | 35,672.03 | 6 | Washington | 25,058.10 |
| 7 | Louisiana | 34,995.58 | 7 | Hawaii | 26,078.61 |
| 8 | South Carolina | 34,465.43 | 8 | District of Columbia | 26,349.25 |
| 9 | Wisconsin | 34,355.14 | 9 | Virginia | 26,513.61 |
| 10 | Mississippi | 34,031.31 | 10 | Idaho | 26,778.19 |

**TABLE N13.** High Gi* Value (Hotspots) and Low Gi* Value (Coldspots) States.

| Rank | Hotspot States | Gi* Value | Rank | Coldspot States | Gi* Value |
| --- | --- | --- | --- | --- | --- |
| 1 | Arkansas | 1.000414 | 1 | Alaska | -1.237323 |
| 2 | Georgia | 1.582820 | 2 | Delaware | -1.139304 |
| 3 | Mississippi | 1.556684 | 3 | New Hampshire | -1.693379 |
| 4 | Missouri | 1.395873 | 4 | Idaho | -1.346255 |
| 5 | Indiana | 1.060011 | 5 | Pennsylvania | -1.133016 |
| 6 | Alabama | 1.032853 | 6 | New Jersey | -1.765067 |
| 7 | Texas | 1.071157 | 7 | New York | -1.178878 |
| 8 | Ohio | 1.126834 | 8 | Vermont | -1.946207 |
|  |  |  | 9 | District of Columbia | -1.987696 |
|  |  |  | 10 | Oregon | -1.348064 |

## Notes

### Competing Interest Statement

The authors have declared no competing interest.

### Funding Statement

This study did not receive any funding

